# Deciphering Risk of Recurrent Bone Stress Injury in Female Runners Using Serum Proteomics Analysis and Predictive Models

**DOI:** 10.1101/2024.12.03.24318372

**Authors:** Genevieve E. Romanowicz, Kristin Popp, Ethan Dinh, Isabella R. Harker, Kelly Leguineche, Julie M. Hughes, Kathryn E. Ackerman, Mary L. Bouxsein, Robert E. Guldberg

**Affiliations:** Knight Campus for Accelerating Scientific Impact, University of Oregon, Eugene, OR; Endocrine Division, Massachusetts General Hospital, Boston, MA; Boston Children’s Hospital, Boston, MA; TRIA Orthopedics, HealthPartners Institute, Bloomington, MN; Military Performance Division, United States Army Research Institute of Environmental Medicine, Natick, Massachusetts, USA; Center for Advanced Orthopaedic Studies, Beth Israel Deaconess Medical Center, Boston, MA

## Abstract

Up to 40% of elite athletes experience bone stress injuries (BSIs), with 20-30% facing reinjury. Early identification of runners at high risk of subsequent BSI could improve prevention strategies. However, the complex etiology and multifactorial risk factors of BSIs makes identifying predictive risk factors challenging. In a study of 30 female recreational athletes with tibial BSIs, 10 experienced additional BSIs over a 1-year period, prompting investigation of systemic biomarkers of subsequent BSIs using aptamer-based proteomic technology. We hypothesized that early proteomic signatures could discriminate runners who experienced subsequent BSIs. 1,500 proteins related to metabolic, immune, and bone healing pathways were examined. Using supervised machine learning and genetic programming methods, we analyzed serum protein signatures over the 1-year monitoring period. Models were also created with clinical metrics, including standard-of-care blood analysis, bone density measures, and health histories. Protein signatures collected within three weeks of BSI diagnosis achieved the greatest separation by sparse partial least squares discriminant analysis (sPLS-DA), clustering single and recurrent BSI individuals with a mean accuracy of 96 ± 0.02%. Genetic programming models independently verified the presence of candidate biomarkers, including fumarylacetoacetase, osteopontin, and trypsin-2, which significantly outperformed clinical metrics. Time-course differential expression analysis highlighted 112 differentially expressed proteins in individuals with additional BSIs. Gene set enrichment analysis mapped these proteins to pathways indicating increased fibrin clot formation and decreased immune signaling in recurrent BSI individuals. These findings provide new insights into biomarkers and dysregulated protein pathways associated with recurrent BSI and may lead to new preventative or therapeutic intervention strategies.

**One Sentence Summary:** Our study identified candidate serum biomarkers to predict subsequent bone stress injuries in female runners, offering new insights for clinical monitoring and interventions.

## INTRODUCTION

Bone stress injuries (BSIs) are common in active individuals, affecting professional and amateur athletes (1 in 20 individuals) *(1, 2)* and military personnel undergoing initial military training (as high as 1 in 10 individuals) *(1, 3, 4)*. For athletes, BSIs are one of the most burdensome injuries, with 20-40% of athletes experiencing BSIs during training *(5, 6)*. While BSIs can range in severity, they may have an outsized financial and psychological impact due to high rates of additional BSIs and extended absences from sport *(7)*. Methods to improve BSI healing, enhance early detection, and inform preventative strategies would remove an obstacle to training and improve athlete well-being. Yet the etiology and pathogenesis of BSIs are multifactorial and complex, making identification of a single factor leading to subsequent BSIs (new BSI at the same or different location) and ultimately the prevention of new BSIs challenging.

A history of a prior BSI is one of the strongest risk factors for a recurrent BSI *(2, 6)*. Diagnosis of these injuries is accomplished through magnetic resonance imaging (MRI), x-ray based imaging, or clinical symptoms. The standard of care for return to play is based off absence of pain with progressive physical activity after an initial period of rest/unloading. For high-risk BSIs, repeat MRI may also be ordered to ensure healing. However, current biomarkers *(8)*, clinical assessments [MRI, dual-energy x-ray absorptiometry (DXA), and computed tomography (CT)] *(9)* lack reliable sensitivity and specificity in identifying individuals at risk of subsequent BSIs. Return to sport is highly individualized based on the patient specific risk factors and pain levels with loading *(10)*. Together, variability in diagnostics, prognostics, risk factors, and treatment regimens has made the identification of individuals at risk of subsequent BSIs challenging.

Although most runners with BSIs are able to return to activity after rest, the frequency of subsequent BSIs ranges from 20-30% *(11, 12)*. Recurrence of BSIs at the same or different locations, are ∼3 times greater in women compared to men *(5)*. Recovery from a BSI can range from 4 weeks to 6 months *(9, 12, 13)*, causing extended removal from sport. While thus far elusive, identification of a reliable biomarker or biomarker panel, would dramatically improve clinical decision making for return to activity and post-injury management as well as prevention and treatment of BSIs.

In our previously reported study, female recreational runners who experienced a subsequent BSI were younger, more likely to have a history of prior skeletal fracture, and had later onset of menses and lower serum parathyroid hormone (PTH) levels than female athletes who did not experience a subsequent BSI *(11)*. Notably, bone density and microarchitecture was largely similar between athletes who did and did not suffer a recurrent BSI, though women who had a reinjury had lower cortical bone tissue mineral density (TMD) and estimated stiffness at the distal tibia *(11)*. Altogether, there was no set of clinical or performance-related factors that distinguished between individuals who would sustain a subsequent BSI during the one year follow up and those who would heal uneventfully. This informed our current study with the objective to identify protein signatures in individuals who experience subsequent BSIs to address these clinical challenges.

The identification of biomarkers in bone healing is an emerging field, supported by the development of sophisticated machine learning (ML) models designed to navigate the complexities of high-dimensional datasets characterized by limited sample sizes, multi-omic integration, and biological noise *(14)*. Genetic programming-based ML approaches have proven effective in extracting critical insights from such datasets. Additionally, high-throughput proteomic technologies, such as the SOMAscan assay *(15–17)*, enable simultaneous quantification of over 1,500 proteins from human serum with exceptional precision, enhancing the detection of biological signals. However, these advancements also amplify the challenge of “small n” datasets, where the number of variables vastly exceeds the sample size, increasing the risk of overfitting *(18)*. Despite this, when strong signals exist, it is possible to identify highly informative feature sets which are predictive in the clinical context.

Our study leverages established machine learning and genetic programming models to interrogate both linear and non-linear relationships within high-resolution proteomic data. By combining advanced proteomic profiling with ML, we provide critical insights into diagnostics, early intervention, and personalized risk assessment in sports medicine. Furthermore, our dataset represents the most comprehensive serum proteomic analysis for BSI studies to date, illuminating dysregulated biological pathways in an underexplored population of female athletes.

## RESULTS

### Study Cohort

Utilizing samples serum from our previous study *(11)*, we sought to characterize the proteomic signatures of individuals with and without additional BSI’s over a one-year recovery period. The original study enrolled female recreational athletes (n=30) (Fig. 1) from the local community who were diagnosed with a tibial BSI with MRI Grading of 2-4/4 by Fredericson Criteria *(11)*. Patients were enrolled within 3 weeks of initial BSI MRI diagnosis. At enrollment, areal bone mineral density (aBMD) of the hip and spine were assessed by DXA, while volumetric bone mineral density (vBMD) and bone microarchitecture at the distal tibia were measured via high-resolution peripheral quantitative computed tomography (HR-pQCT). HR-pQCT scans were also collected at each follow up visit, including 6, 12, 24, and 52 weeks after the initial enrollment. Additionally, clinical metrics were collected, including laboratory blood values, pain assessments, physical activity assessments, menstrual status, and health and fracture history (full list in Data File S1, data previously reported *(11)*). Of the 30-runner cohort, 10 experienced additional BSIs at times ranging from 6-52 weeks after initial diagnosis *(11)*. The majority occurred between weeks 12 and 52 (9 out of 10 additional BSIs) (Table 1). One individual had a third BSI event in the 24-to-52-week period. Only one additional BSI was in the same location as the first BSI. For all individuals, serum was analyzed retrospectively for protein signatures of additional BSI (full list in Data file S2).

**Figure 1:**
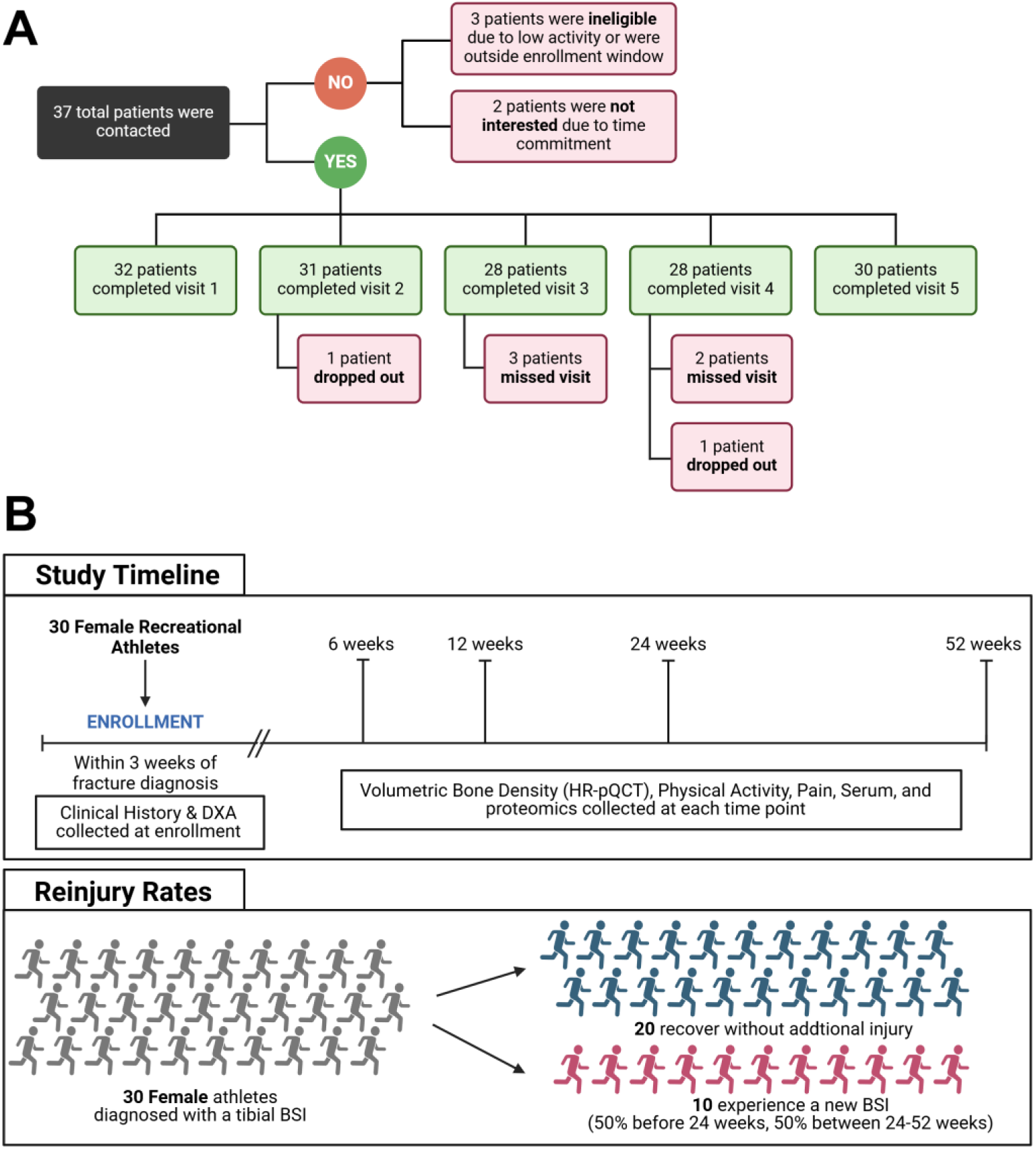
Study design and patient attrition. (A) Female recreational athletes were recruited to participate in an observational study following bone stress injury (BSI) diagnosis with an MRI Fredericson grade 2-4/4. Patient attrition and visit completion are noted. *Figure adapted from Popp, et al. 2019.* (B) All patients were enrolled within 3 weeks of diagnosis of a tibial BSI and monitored for metrics such as bone morphometry changes, physical activity, and pain. Serum was collected and originally analyzed for standard of care laboratory values. This study re-analyzed the serum for 1,500 proteins across all individuals and times. Of the 30 individuals, 20 went on to recover without additional injury. 10 individuals experienced a second BSI, with one individual experiencing a third BSI over the one year follow up (Table 1). Created in https://BioRender.com

**Table 1.** Summary of additional BSI events during the 52 weeks of monitoring. A total of 10 of 30 participants experienced a 2^nd^ BSI, with one individual experiencing a 3^rd^ BSI. Only one BSI was at the same location as the original BSI. Data previously reported in *Popp, et al., 2019*.

| Time Window | 2<sup>nd</sup> BSI | 3<sup>rd</sup> BSI |
| --- | --- | --- |
| Enrollment - 6 weeks | 1 |  |
| 6 - 12 weeks | 0 |  |
| 12-24 weeks | 5 |  |
| 24-52 weeks | 4 | 1 |
| **Total:** | 10 Additional BSIs |  |

### Additional and single BSI groups can be identified based on early protein signatures at time of enrollment and before any additional BSI events

The sparse partial least squares discriminant analysis sPLS-DA model was chosen to identify key features from the enrollment visit (visit 1, < 3 weeks after initial BSI diagnosis), separating additional BSI individuals from single BSI individuals. Models were first applied to the clinical outcomes *(11)*. Separation was not clear for the clinical data, with the top-ten features identified as age at menses onset, cortical TMD (Ct. TMD), compressive stiffness (Comp. Stiff.), age, failure load (F. Load), and prior number of fractures (Prior FX) (Fig. 2A&D), all of which were previously reported *(11)* to be different between additional and single BSI groups. Serum procollagen type I N-propeptide (P1NP) and serum osteocalcin were also included in the top-ten features (Fig. 2D, Data File S3). These serum metrics were measured using standard-of-care clinical laboratory assays in the previous study *(11)*, and are different than the aptamer based serum proteomic data. Due to the small sample size, feature importance estimation via bootstrapping was conducted on the sPLS-DA models. The bootstrapping method included repeated removal of one participant and recording of the top model features. Stable features, identified as those appearing in at least 80% of models following bootstrapping (Data File S4), were then evaluated for predictive performance via Receiver Operating Characteristic (ROC) analysis. Due to the small sample size and class imbalance in the data, leave one out cross validation (LOOCV) was applied. For the clinical data, the features selected were medical history (age, age at menses onset, prior history of fracture), tibia morphology via HR-pQCT (CtTMD, stiffness and strength), and serum markers of bone metabolism (P1NP, tartrate-resistant acid phosphatase 5b (TRAP5b)). The area under the curve (AUC) for mean ROC curve indicated an average accuracy of 79% ± 0.08 across various predictive models including Logistic Regression, Random Forest, and CatBoost (Fig. 2G). The average sensitivity was 63.3% and average specificity was 76.7% (Fig. S1A). The positive predictive value was 57.6% and negative predictive value was 80.7% (Fig. S1A).

**Figure 2:**
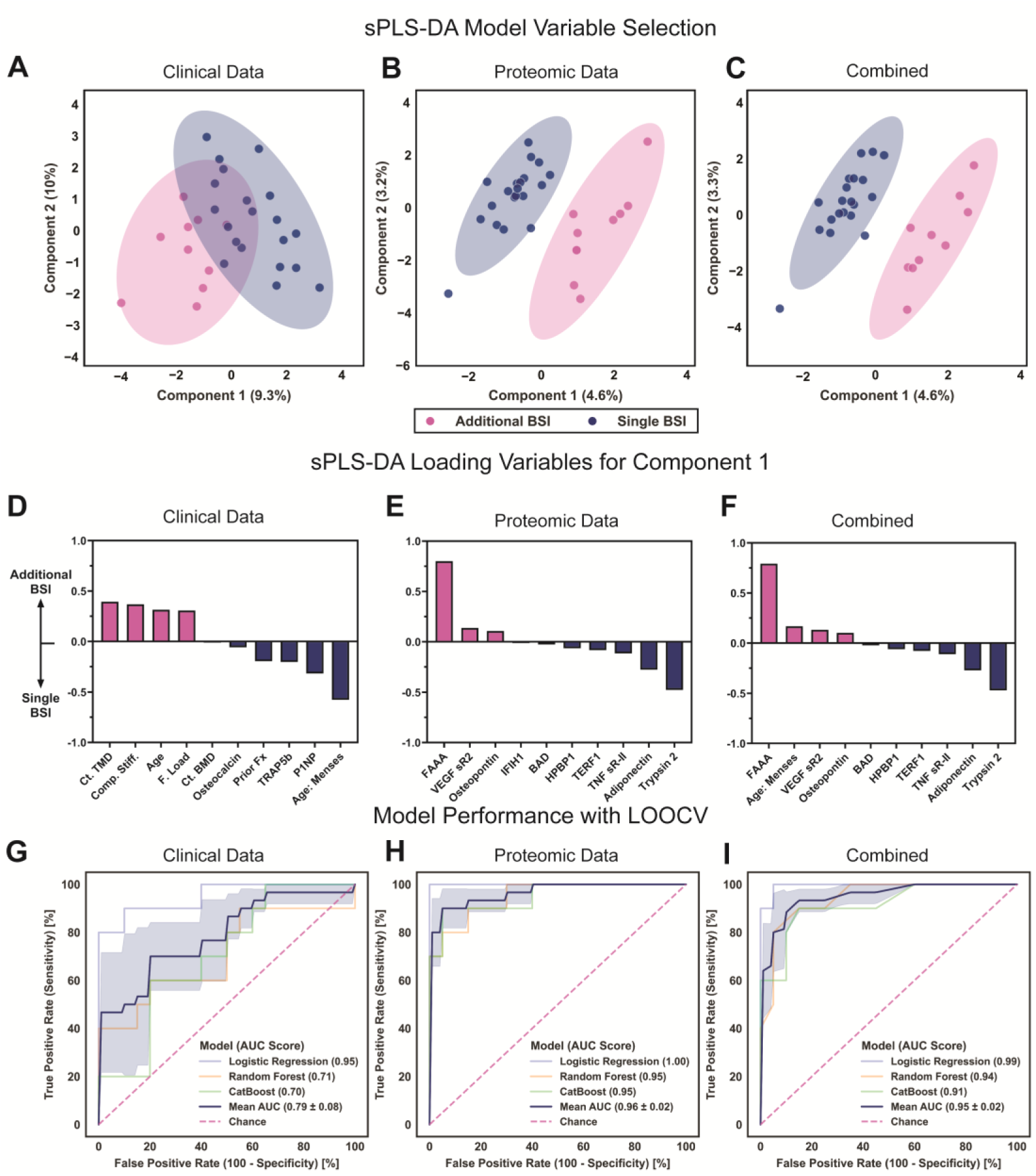
Bone stress injury (BSI) reinjury classification can be identified from proteomic data within 3 weeks of initial BSI. (A) Clinical data were used to separate additional from single BSI individuals with a sparse partial least squares discriminant analysis (sPLS-DA) model but yielded no clear separation. (B) Proteomic signatures were then tested in a similar model, increasing separation. (C) Combining both clinical and proteomic data marginally improves separation. (D) The top latent variables for the sPLS-DA model are shown for clinical data, (E) proteomic data, and (F) the combined clinical and proteomic data. (G) The most frequent variables identified by feature importance estimation via bootstrapping were input into the sPLS-DA models and tested in predictive models, including Logistic Regression, Random Forest, and CatBoost, each with leave one out cross validation (LOOCV). Receiver operating characteristic (ROC) curves are shown with a mean accuracy of 79% ± 0.08 for clinical data, (H) 96% ± 0.02 for proteomic data, and (I) 95% ± 0.02 for the combined data. Shaded regions (A-C) represent the 95% confidence interval. AUC values are mean ± S.E.

Similar models were generated for the serum proteomic data. sPLS-DA was performed on serum proteomic data from the enrollment visit. Ten proteins were identified which segregated the single vs. additional BSI groups (Fig. 2B). Of these, fumarylacetoacetase (FAAA), trypsin-2, and adiponectin were identified with the highest coefficients and greatest influence on the models to discriminate between single and additional BSIs (Fig. 2E). Less influential, but still important features included soluble vascular endothelial growth factor receptor 2 (VEGF sR2), tumor necrosis factor (TNF) receptor superfamily member 1B (TNF sR-II), osteopontin (OPN), telomeric repeat binding factor 1 (TERF1), Hsp70-binding protein 1 (HPBPI), Bcl2-associated agonist of cell death (BAD), and Interferon-induced helicase C domain-containing protein 1 (IFIH1) (Data File S3). Features present in at least 80% of models were FAAA, tyrpsin-2, adiponectin, OPN, TNF sR-II, VEGF sR2 (Data File S4). These proteins were then evaluated with an ROC analysis and LOOCV. The AUC for the mean ROC curve indicated an average accuracy of 96% ± 0.02 across various predictive models including Logistic Regression, Random Forest, and CatBoost (Fig. 2H). The average sensitivity was 80.0% and average specificity was 98.3% (Fig. S1B). The positive predictive value was 96.0% and negative predictive value was 90.8% Fig. S1B). The proteomic data models performed better (mean AUC ± S.E.) than clinical data (*p* = 0.012, one-way ANOVA).

To determine if combining datasets would improve accuracy of the models, we then repeated the sPLS-DA models with both the clinical and the proteomic datasets. Combined datasets slightly increased separation between the two patient populations (Fig. 2C). Yet, of the top-10 variables, only one clinical metric (age at onset of menses) was included (Fig. 2F, Data File S3). Features present in at least 80% of models included FAAA, trypsin 2, age at menses onset, adiponectin, TNF sR-II, and OPN (Data File S4). These six features were then evaluated with a ROC analysis and LOOCV. The AUC for the mean ROC curve indicated an average accuracy of 95% ± 0.02 across various predictive models including Logistic Regression, Random Forest, and CatBoost (Fig. 2I). The average sensitivity was 80.0% and average specificity was 93.3% (Fig. S1C). The positive predictive value was 85.7% and the negative predictive value was 90.3% (Fig. S1C). While the average accuracy was similar between the proteomic data model and the combined data model, the combined models had a small reduction in sensitivity with increased specificity. The combined models performed better (mean AUC ± S.E.) than clinical data (*p* = 0.017, one-way ANOVA), but not better than proteomic data (*p* = 0.97, one-way ANOVA).

### Genetic programming models further support identification of candidate biomarkers of subsequent BSIs

Genetic programming has become a useful tool to generate predictive models in biological datasets where both linear and non-linear relationships may occur simultaneously *(19, 20)*. We thus explored if linear and non-linear models would improve the predictive accuracy of both the proteomic and clinical datasets using Evolved Analytics DataModeler software. Thousands of models are generated across at least 30 successive generations, in which “fit” models are selectively bred. Finally, models are rated based on accuracy and complexity, and those with optimal complexity/accuracy tradeoffs are aggregated into a model ensemble. Complexity refers to a measure of numerical operations required to fit the data into either the additional or single BSI group. Lower complexity is desirable, with a complexity score > 80 considered too complex for our dataset based on sample size. Therefore, we sought to evaluate the complexity and accuracy of produced models for each of our datasets. Resultant model ensembles were used to predict a binary outcome measure: single vs. additional BSI. This approach allowed for an independent statistical validation of candidate biomarkers by utilizing a completely independent machine learning approach as compared to the linear models used in sPLS-DA. An additional advantage of genetic programing is that candidate equations that are used to generate the model ensembles are displayed as an output, showing the algorithmic relationship between features and enhancing interpretability.

Datasets were compared for accuracy and error including ‘Clinical Data, ‘Proteomic Data’, or ‘Combined Data’ with the same datasets tested in the sPLS-DA models. Ten candidate models were selected from the thousands of models generated per category. Candidate models were restricted to 4 variables, which is the recommended number of variables to allow for at least a 5:1 ratio of datapoints (individuals) to support selected variables (Fig. 3A-C). The first step in evaluating genetic programming model ensembles (collections of models) is to inspect if model ensembles exhibit a ‘knee’ shaped Pareto plot, which signifies a balance between minimizing error and avoiding unnecessary complexity. Minimizing error with balance complexity is a desirable trait in all models but is especially important with small datasets. The ‘knee’ shape was notably absent in models relying solely on clinical data (Fig. 3A) suggesting that accuracy could not be achieved without undue complexity and therefore risk of overfitting. Additionally, accuracy in this context signifies confidence in a binary classification (e.g., single or additional BSI), rather than estimate of a continuous variable.

**Figure 3:**
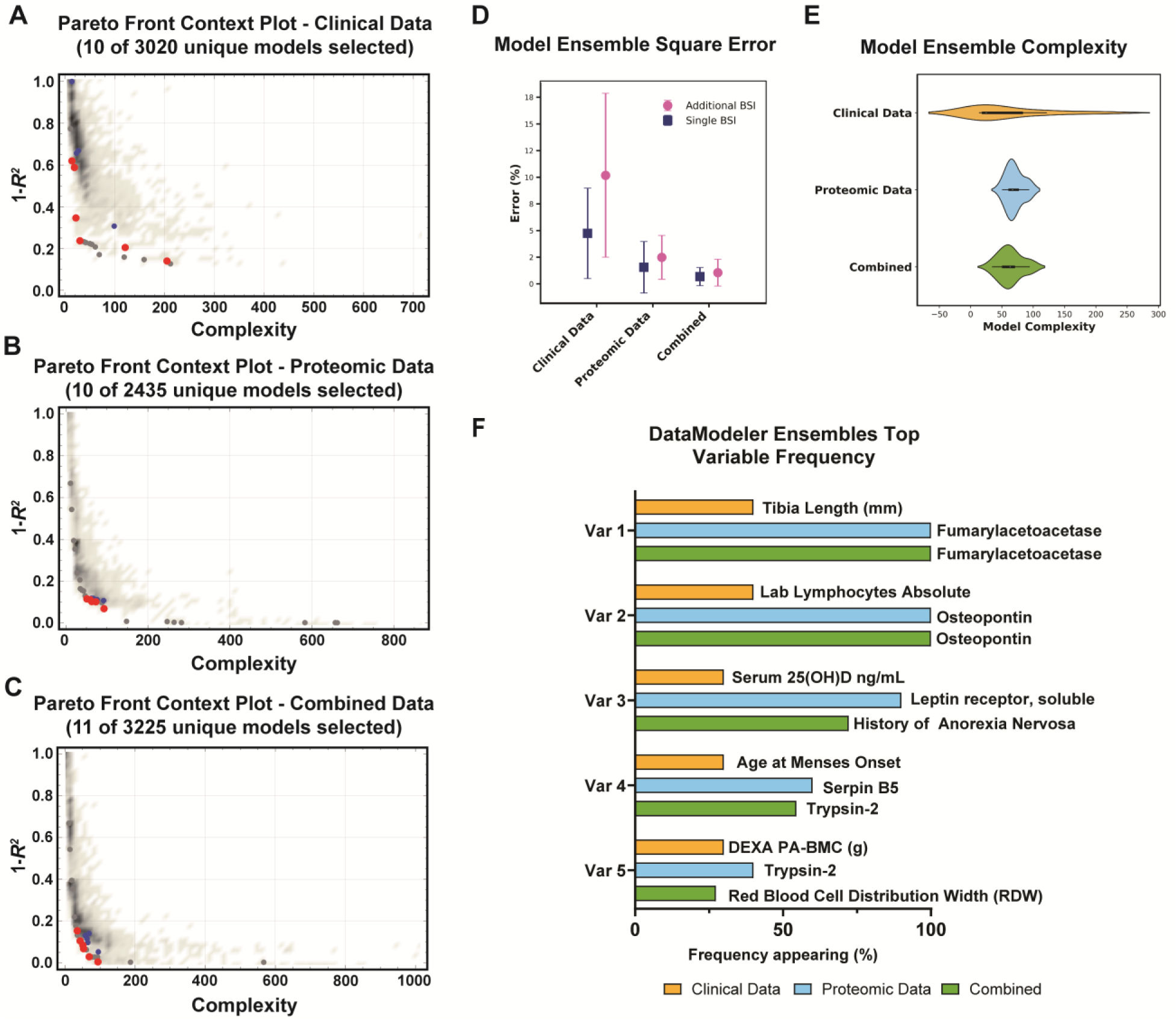
Genetic programming models were evaluated for accuracy and complexity using clinical, proteomic, and combined datasets. Models were generated using genetic programming to create candidate models and evaluate complexity and accuracy. The top ten models are highlighted in red and blue circles. (A) Clinical data produced minimal models that were both accurate and complex, noted by the absences of the ‘knee’ configuration in the plot. (B) Proteomic data produced the characteristic ‘knee’ shape, allowing for exploration of features that generate highly accurate and minimally complex models. (E) Model accuracy was compared as well as (F) model complexity, with combined datasets performing best. (D) Variable presence in the model ensembles were compared between clinical, proteomic, and combined datasets. Clinical data lacked variables present in >50% of the generate models. In contrast, proteomic data produced 4 variables present in > 50% of models with FAAA appearing in 100% of models. Combined datasets performed similarly with the inclusion of a history of anorexia as the top feature from the clinical dataset.

Genetic programming models for the clinical metrics had the poorest performance, with resultant 10-model ensemble (Table S1) yielding relatively low accuracy (R^2^ = 78%) and high error rates for single BSI predictions (17 ± 14%) and for additional BSI predictions (29 ± 13%) across the selected 10 best performing models (Fig. 3D&E). Substantial variability between model ensembles was also present in clinical metrics alone (Fig. 3D). For the clinical models, the top-5 most frequently occurring variables across the model ensemble included tibial length (40%), absolute lymphocyte count (40%), age at menses onset (40%), serum levels of 25 (OH) vitamin D (30%), and posteroanterior lumbar spine bone mineral content (PA-BMC) by DXA (30%). Furthermore, no variable was present in more than 50% of the selected models (Fig. 3F). The complexity scores ranged from 35-205, achieving 15.2-80% accuracy, respectively. A complexity score of 205 is extremely high and suggestive of overfitting. This ultimately led to highly complex models with considerable variability (Fig. 3E). Most of the resulting models were linear, with subsets of models being non-linear for the variables PA-BMD and PA-BMC (Fig. S2)

These results are in contrast with the model ensemble using proteomic data, which demonstrated higher accuracy (R^2^ = 91.6%), mean complexity score of 70.3, and much lower average error rates for single BSI predictions (3 ± 8%) and for additional BSI predictions (8 ± 14%) across the selected 10 best performing models (Fig. 3D&E, Table S2). The expected ‘knee’ shape is present (Fig. 3B) in the proteomic models, allowing for exploration of a variety of models with balanced accuracy and complexity. For the proteomic models, the top-5 most frequently occurring variables across the models included FAAA (100%), OPN (100%), soluble leptin receptor (LEPR) (90%), Serpin B5 (60%), and trypsin-2 (40%) (Fig. 3F). From the variable distribution analysis of 2,435 generated models, a frequency of 60-100% in the resultant model ensemble underscores the predictiveness and consistency of FAAA, OPN, LEPR, and Serpin B5 compared to clinical data, which did not have a single variable represented in more than 50% of models. Resultant models were primarily linear with non-linear models for OPN and LEPR (Fig. S3)

Combining both clinical and proteomic data yielded the highest accuracy (R^2^ = 97%) and lowest error rates for single BSI predictions (3 ± 8%) and for additional BSI predictions (5 ± 9%), with a modest mean complexity score (63.2) across the selected 11 best performing models (Fig. 3D&E, Table S3). Again, the ‘knee’ shape is present in the model ensembles (Fig. 3C). When clinical and proteomic data were combined, history of anorexia replaced LEPR as the third most common frequent variable (67% of models) in the combined ensembles, yet history of anorexia had not previously been identified in the clinical data models (Fig. 3F). Of note, only two individuals had a history of anorexia nervosa. For the combined models, the top-5 most frequently occurring variables across the selected models included FAAA (100%), OPN (100%), history of anorexia nervosa (72.7%), trypsin-2 (54.5%), red blood cell distribution width (RDW) (27.3%) (Fig. 3F). Resultant models were primarily linear with non-linear models for FAAA and OPN (Fig. S4)

### Candidate biomarkers have a temporal expression pattern in additional vs. single BSI individuals

The top protein features from the sPLS-DA and genetic programming models were analyzed over the one-year follow-up including baseline, 6, 12, 24, and 52 weeks. Exploring candidate biomarker proteins across sPLS-DA models and genetic programming, we investigated the temporal patterns of FAAA, trypsin-2, and osteopontin. FAAA levels were higher in individuals who experienced additional BSIs compared to those with a single BSI at enrollment, week 6, and week 24 (enrollment, *p* = 0.004; week 6, *p* = 0.03; week 24, *p* = 0.05) (Fig. 4A). By contrast, Trypsin-2 levels were consistently lower in individuals with additional BSIs compared to those with a single BSI at all examined timepoints (enrollment, *p* = 0.002; week 6, *p* = 0.02; week 12, *p* = 0.003; week 24, *p* = 0.01; week 52, *p* = 0.003) (Fig. 4B). OPN levels were higher in individuals with additional BSIs compared to those with a single BSI at enrollment and week 24 (enrollment, *p* = 0.01; week 24, *p* = 0.005) (Fig. 4B). Given that the occurrence of additional BSIs varied in timing among individuals, we were unable to identify the FAAA, trypsin-2, or OPN levels that corresponded to the timing of the second or third BSI onset.

**Figure 4:**
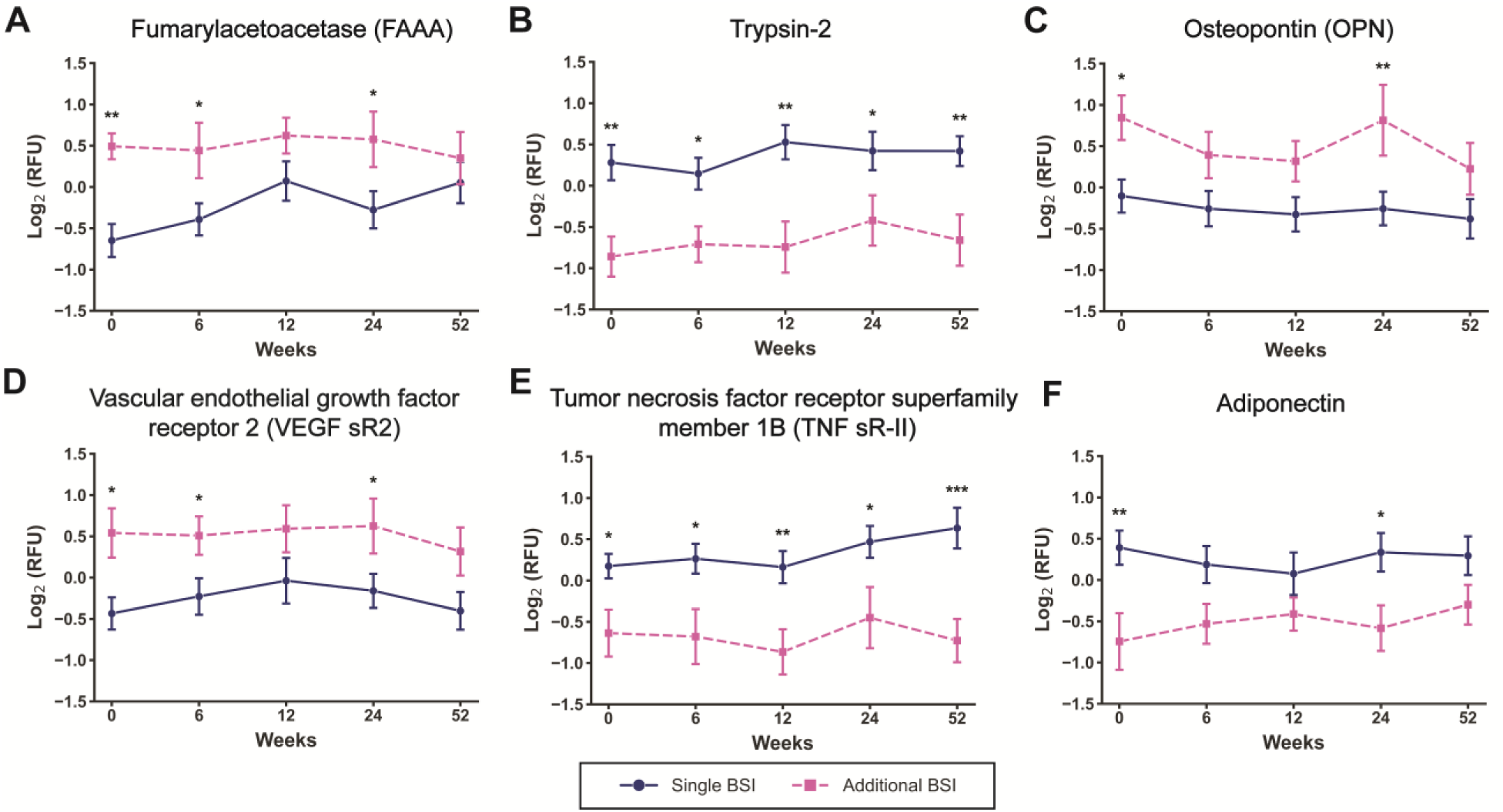
Serum proteins represented in predictive models displayed distinct expression patterns over time between additional and single bone stress injury (BSI) groups. (A-C) Proteins identified in both sparse partial least squares discriminant analysis (sPLS-DA) and genetic programming models, such as fumarylacetoacetase (FAAA), trypsin-2, and osteopontin (OPN), exhibited temporal expression differences. (D-F) Proteins identified only in the sPLS-DA model, including vascular endothelial growth factor receptor 2 (VEGF sR2), tumor necrosis factor receptor superfamily member 1B (TNF sR-II), and adiponectin, also showed significant temporal variation. Data are presented as mean ± SEM (n = 10 for additional BSI, n = 20 for single BSI, analyzed using linear mixed-effects model (\**p* < 0.05, \*\**p* < 0.001).

We further explored the time dependent differences in key features from the sPLS-DA models, specifically characterizing VEGF sR2, TNF sR-II, and adiponectin (Fig. D-F). VEGF sR2 levels were higher in individuals who experienced additional BSIs compared to those with a single BSI at enrollment, week 6, and week 24 (enrollment, *p* = 0.01; week 6, *p* = 0.04; week 24, *p* = 0.05) (Fig. 4D). By contrast, TNF sRII levels were consistently lower in individuals with additional BSIs compared to those with a single BSI at all examined timepoints (enrollment, *p* = 0.02; week 6, *p* = 0.01; week 12, *p* = 0.003; week 24, *p* = 0.01; week 52, *p* = 0.0002) (Fig. 4E). Adiponectin levels were lower in individuals with additional BSIs compared to those with a single BSI at enrollment and week 24 (enrollment, *p* = 0.004; week 24, *p* = 0.05) (Fig. 4F). Again, the protein levels were most different between groups at enrollment and at week 24, immediately following 3 additional BSIs events (Table 1). Yet, proteomic expression of FAAA and Trypsin-2 showed sustained temporal patterns in proteomic levels between single and additional BSI groups (Fig. 4A&B). Notably, week 24 was different for FAAA, trypsin-2 and OPN, which followed 5 of the additional BSI events and preceded 5 more BSIs (Table 1), suggesting that these proteins may be elevated following or prior to subsequent BSI.

### Additional BSI was associated with early dysregulation in proteins involved in blood clotting and immune function

While proteomic analysis was useful for the biomarker identification and risk classification, biomarkers alone do not give insights into biological pathways involved in additional BSI risk. Therefore, functional pathways altered between additional and single BSI groups were assessed through a gene set enrichment analysis *(21)*. Proteins were first analyzed for differential expression between groups while blocking for time using the ExpressAnalyst package *(22)* based on the *limma* package *(23)* in R (R 4.4.1). This analysis revealed 112 significantly different proteins expressed between additional and single BSI individuals [log2 FC > 0.5, Benjamini-Hochberg false discovery rate (FDR) adjusted p-value < 0.05] (Data File S5). Notably, 76 of these genes were found to be upregulated, while 36 were downregulated, indicating substantial biological divergence that corresponds to subsequent BSIs (Fig. 5A). Of these proteins, several key proteins identified in sPLS-DA and genetic programming models were also significantly differentially expressed proteins, including FAAA, trypsin-2, OPN, TNFs RII, VEGF sR2, LEPR, and adiponectin (Fig. 5A).

**Figure 5:**
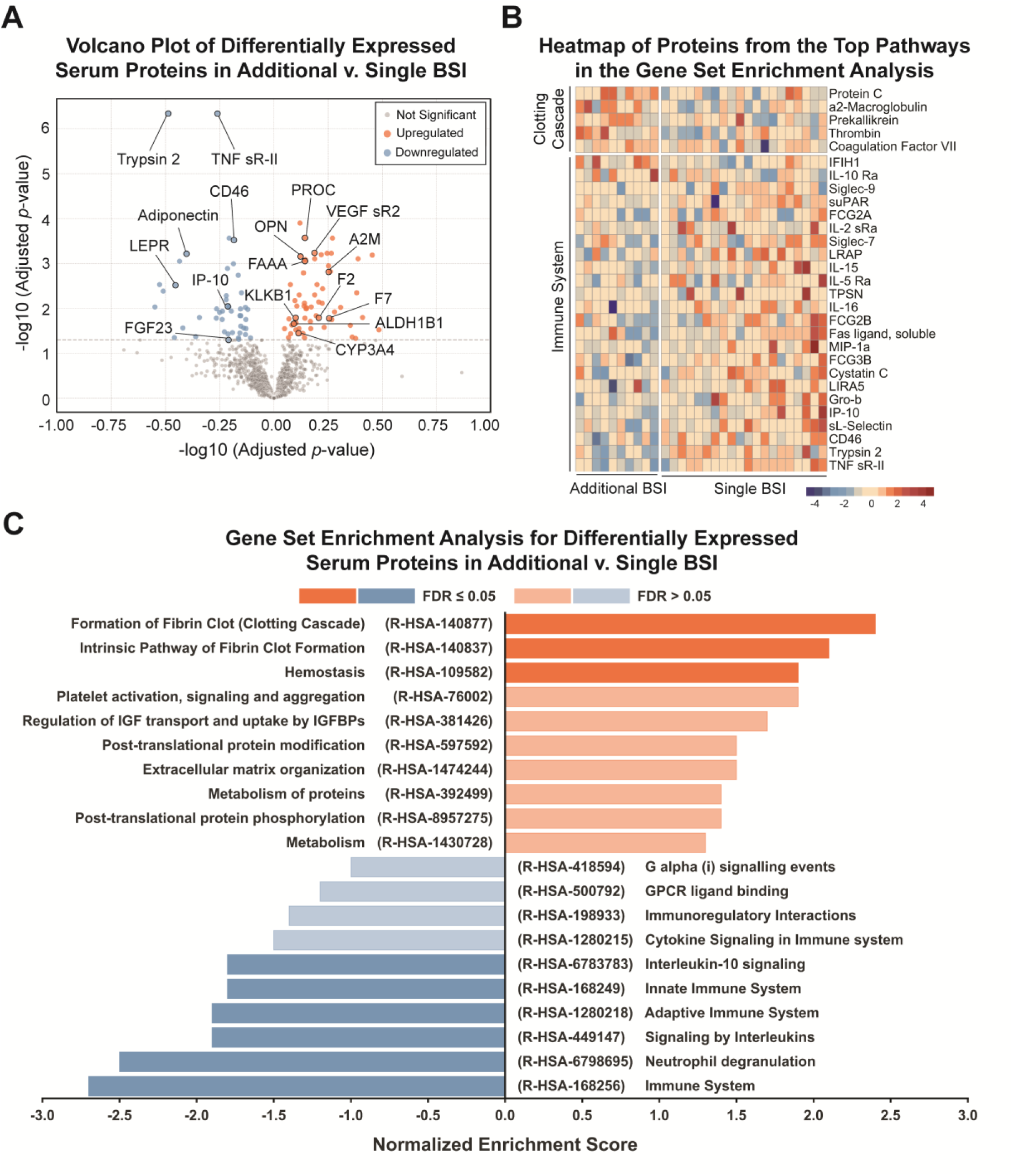
Individuals with additional bone stress injury (BSI) show increased blood clotting and decreased immune system proteins over time. (A) Volcano plot of differentially expressed serum proteins up- or downregulated in additional BSI v. single BSI individuals. (B) Heatmap of proteins from pathways including “formation of fibrin clot” and “immune system” that were identified in (C) a gene set enrichment analysis of differentially expressed proteins in additional BSI v. single BSI individuals. All data are aggregated per individual over 52 weeks and analyzed, blocking for time, for both the differential expression and gene set enrichment analysis (GSEA) analysis.

To enable Gene Set Enrichment Analysis (GSEA) with proteins, the initial panel of 1,500 proteins were mapped to 1,207 corresponding unique genes via EntrezID to enable the analysis through established gene network databases. Dysregulated pathways were identified in individuals with an additional BSI as compared to a single BSI. Downregulated pathways in the additional v. single BSI groups included immune system function, adaptive and innate immunity, interleukin-10 (IL-10) signaling, neutrophil degranulation, and signaling by interleukins (Fig. 5C). Significantly upregulated pathways in the additional v. single BSI included formation of fibrin clot, intrinsic pathway of fibrin clot formation, and hemostasis (Fig. 5C). A full list can be found in the supplement (Data File S6). Of the significant pathways, six are primarily associated with the immune system, while the remaining three are linked to blood clotting processes. Formation of fibrin clot (clotting cascade) (R-HSA-140877) included protein C (PROC), a2-macroglobulin (A2M), kallikrein B1 (KLKB1), thrombin (F2), and coagulation factor VII (F7) (Fig. 5B). Immune system (R-HAS-168256) included 24 immune system related proteins, notably trypsin-2 and TNF sRII, which were present in the sPLS-DA models (Fig. 5B).

Because blood clotting pathways were significantly upregulated in additional BSI individuals, we explored if type of contraceptive (combined oral, progestin-releasing IUD, previously history of combined oral, or never used) contained any overlap with the protein profile identified in additional BSI individuals. Hormonal contraceptives may influence blood clotting pathways *(24)* as well as bone density and metabolism *(25)* and may be a confounding factor in this study. However, our sPLS-DA analysis of protein signatures relating to type of contraceptive did not contain overlapping proteins with the dysregulated proteins in the additional BSI individuals (Fig. S4 A&C). We also explored if MRI grade was associated with the dysregulated proteins in this study but found no overlap in MRI grade proteins and BSI risk proteins (Fig. S4 B&D).

## DISCUSSION

Pathogenesis of BSIs is both complex and remains poorly understood. Biological and biomechanical factors contribute to subsequent BSIs, but clear clinical risk indicators are lacking. This study aimed to identify protein signatures in individuals who experience subsequent BSIs to address these clinical challenges. We conducted an in-depth proteomic analysis of female athletes with and without recurrent BSI. Our findings revealed that subsequent BSI risk can be predicted by early serum proteomic profiles. Using multivariate modeling, a sparse partial least squares discriminant analysis (sPLS-DA) model identified six candidate biomarkers (FAAA, trypsin-2, OPN, TNF sR-II, and VEGF sR2) with an accuracy of 91-96% in predicting recurrent BSI risk category from enrollment proteins alone. Analysis of protein changes over time revealed 112 significantly different proteins, mapped to biological pathways associated with decreased immune system function and increased blood clotting pathways.

Genetic programming further validated the candidate biomarkers, identifying FAAA as a key predictor of subsequent BSIs. Notably, clinical metrics alone were insufficient to distinguish between single and recurrent BSI groups. While combining clinical metrics with proteomic data slightly enhanced genetic programming models, combined data did not outperform proteomic data in sPLS-DA models, demonstrating the superior predictive power of proteomic information. This integration of high-resolution proteomic data enabled the application of standard machine learning methods on a relatively small dataset, surfacing a highly informative feature set far surpassing the predictive value of standard clinical data. Our study not only provides the most comprehensive proteomic dataset for BSIs in the underexplored population of female athletes but also identifies candidate biomarkers for recurrent BSIs, offering valuable insights for targeted therapeutic interventions and improved clinical prediction strategies.

Multivariate linear regression models through sPLS-DA identified FAAA as the marker (at enrollment, < 3 weeks of initial BSI) that most positively correlated with additional BSI and Trypsin-2 as the marker (at all time points) most negatively correlated. These proteins have not been previously identified as biomarkers for BSI risk, and their role in bone health is not well understood. FAAA has been implicated as a serum biomarker for idiosyncratic drug-induced liver injury when increased *(26, 27)* and as a urine biomarker for acute hypercoagulable states in preclinical models when decreased *(28, 29)*. In drug-induced liver inflammation, FAAA positively correlates with alanine aminotransferase (ALT) *(26)*. While ALT was not assessed in our clinical metrics, ALT was measured via our Somamer panel and was found to positively correlate with FAAA levels (Fig. S5, Adj. R^2^ = 0.19, *p* < 0.0001), yet no overt liver diseases were reported in patient medical histories. Alcohol consumption was also not evaluated in this study. Participants were taking a variety of medications, precluding any clear correlations with potential drug-induced liver injury. Another consideration is the use of athletic or herbal supplements, which can be associated with liver injury *(30)* but are difficult to track due to poor reporting and labeling. Although we did not adequately assess supplement intake in this study cohort, other studies have shown that runners with a history of BSI report more frequent supplement intake *(31)* and athletes generally take more types of supplements than non-athletes, with elite athletes having the highest rate of supplement use *(32, 33)*. In the additional BSI group, the proteins FAAA, aldehyde dehydrogenase 1 family member B1 (ALDH1B1), and cytochrome P450 family 3 subfamily A member 4 (CYP3A4) were all upregulated together and relate to the functional pathway of “metabolism” (R-HSA-1430728). Both ALDH1B1 and CYP3A4 are liver enzymes that regulate drug metabolism *(34)* and alcohol metabolism*(35)*. Therefore, elevated liver metabolism enzymes such as FAAA, ALDH1B1, and CYP3A4 suggest a potential challenge to the liver that may be due to supplement use or alcohol intake. Liver health status should be further considered for those at risk of recurrent BSI.

Trypsin-2, also known as PRSS2 or tumor-associated trypsin, has been implicated as a biomarker in pancreatitis *(36)*, ulcerative colitis *(37)*, and diabetes *(38)*, as well as the progression of tumor growth by immunosuppression via interaction with myeloid cells *(39)*. Only recently has trypsin-2 been described in human bone marrow, specifically in the hematopoietic stem cell niche and highly expressed in early, undifferentiated hematopoietic progenitor cells and mobilized CD34^+^ hematopoietic cells *(40)*. However, understanding of the role of trypsin-2 in the bone marrow is limited to the maintenance of the stem cell niche by complex diffusion interaction in the local microenvironments *(40)*, not serum accumulation and potential endocrine signaling within bone marrow. Here we observed that trypsin-2 levels were the strongest positive correlative variable in the sPLS-DA models and the fourth most frequent variable in genetic program models of combined proteomic and clinical data. Trypsin-2 was increased in the single BSI group compared to the additional BSI group at all time points. Only trypsin-2 and TNF sRII were found to be different at all timepoints in the additional BSI group. The diminished trypsin-2 in the additional BSI group corresponds with a reduction in immune pathways, including neutrophil degranulation and innate immune system function, both of which contain trypsin-2 within the pathway categories. These data highlight that elevated serum trypsin-2 levels are positively correlated with successful BSI recovery at early time points and throughout the time course of healing. Diminished trypsin-2 was accompanied by immunosuppressive pathways, suggesting a role for diminished innate immune signaling in those with additional BSIs. A limitation of the interpretation of the absolute levels of trypsin-2 is that this study did not include an uninjured control group, so we cannot determine how the level of trypsin-2 relates to other health conditions in which it has been identified as a biomarker.

Osteopontin (OPN) was identified as the second most frequent variable in genetic programming models for both the proteomic and combined models, occurring in 100% of the models selected in the model ensemble. Yet, OPN was only a modest negative predictor of additional BSI in the sPLS-DA models. OPN also did not appear in any of the differentially expressed pathways but was increased in additional BSI individuals at enrollment and at 24 weeks post injury. OPN was found to be elevated in severe liver dysfunction *(41)*, as well as with severe inflammatory injury and sepsis *(42)*. Elevated levels of OPN are associated with unfavorable prognoses, including mortality, in critically ill patients with SARS-CoV-2 (COVID19) *(43)*. Broadly, OPN is known to have an inflammatory role, and is associated with other pro-inflammatory cytokines, such as C-reactive protein (CRP) and interleukin-6 (IL-6), all of which have been shown to increase following bone trauma *(44)*. We also observed in our time course analysis that OPN peaked in the additional BSI group both at the enrollment visit (within 3 weeks of BSI diagnosis) and again at week 24. This OPN 24-week peak was after an additional BSI in 5 individuals and preceded an additional BSI in 5 others. The elevated levels of OPN and other proteins, at both baseline and week 24 (in and around the additional BSI events), strengthens the evidence that such proteins correspond to injury-related biomarkers. Overall, elevated OPN in the additional BSI group suggests a chronic inflammatory condition.

While immunosuppressive markers are known to correlate to increased fracture risk, this is primarily understood for comorbidities such as organ transplantation and steroid use *(45)*. Similar mechanisms in otherwise healthy populations have not been well studied. However, intense exercise and overtraining can cause a diminished immune response *(46)* and intense exercise produces a robust inflammatory response *(47)*, highlighting a knowledge gap in the role of the immune system in bone quality in athletes. Our pathway analysis for proteins up- or down-regulated with additional BSI risk indicates an impaired or suppressed immune response, which may delay or complicate the recovery process following an injury. In multiple patient studies involving traumatic bone injuries, sustained immune suppression has corresponded to impaired bone healing *(48)*. Of note, IP-10 (also known as C-X-C motif chemokine 10 (CXCL10)), was decreased in the additional BSI group in our time-independent analysis. We have previously shown that serum levels of IP-10 correlated with poor bone healing outcomes in preclinical models of severe muscle and bone trauma with cellular evidence of prolonged immunosuppression *(14)*.

We observed biologically upregulated pathways related to fibrin clot formation, the intrinsic clotting cascade, and hemostasis. Intense exercise is known to induce both coagulation and fibrinolysis, a process typically balanced under normal conditions *(49)*. In our study, we noted an increase in thrombin (F2), a key regulator of hemostasis *(50)*. This increase was not associated with the severity of fractures, as thrombin was not associated with the sPLS-DA for MRI grade (Fig. S4 B&D). While oral contraceptives are thought to create a clinically prothrombotic state, affecting both coagulation and fibrinolysis, we found no association between contraceptive use and blood clotting proteins (Fig. S4 A&C). Elevated proteins within the fibrin clot formation pathways included intrinsic clotting factors, such as KLKB1 and PROC, and extrinsic clotting factors, including coagulation F7, F2, and A2M. The pro-coagulation proteins were elevated alongside inhibitory proteins. Therefore, without functional coagulation assessment, our conclusions are limited to the elevation of proteins attributed to blood clot formation and hemostasis pathways. These findings are consistent with other studies of athletes undergoing intense exercise training *(47, 49, 51, 52)*. In the present study, we did not observe differences in training level between the single and additional BSI groups *(11)*. Therefore, elevated pro-coagulatory proteins suggest that the additional BSI group might be more affected by exercise-related coagulation changes, thereby increasing their risk of subsequent BSIs.

Surprisingly, none of the clinical bone morphometric data were determined to be frequent variables in our combined clinical and proteomic models corresponding to additional BSI risk. Variables appear more frequently in the model ensembles if the features hold higher influence on predictive models and provide generalizability to the larger dataset. In the original study, we observed a decrease in cortical TMD and a decrease in finite element model analysis (FEA) stiffness from the distal tibia in the additional vs. single BSI groups *(11)*. Tibial cortical TMD appeared in the sPLS-DA clinical models as well as tibial cortical vBMD, both of which were acquired using HR-pQCT. Whereas in genetic programming models, the DXA-based lumbar spine BMC appeared in the clinical model ensemble. Interestingly, in the combined model ensemble, DXA-based BMC was replaced by RDW, which has previously been negatively correlated to DXA-based BMD and increased risk of fracture *(53, 54)*. Neither the genetic programming nor sPLS-DA clinical models were highly predictive for clinical data alone. Tibial length was included as the most frequent variable in the genetic program models of clinical metrics alone, but frequency in the models was 40% and comparable to frequency of other top variables, including absolute lymphocyte count (40%), vitamin D (30%), and age at menses onset (30%). Together, this implies that the clinical data variable are present in a relatively flat data plane are likely overfitted in our relatively small dataset. We would expect that if the clinical models were tested on a new dataset, they may not perform well in stark contrast to the proteomic datasets.

When proteomic and clinical data were combined, history of anorexia nervosa emerged as the third most frequent variable, present in 72.7% of the models, replacing LEPR. This finding suggests that both a history of anorexia nervosa and LEPR protein levels may be interchangeable in our models in relation to BSI outcomes. Low leptin levels are consistently noted in those with low weight anorexia nervosa *(55–57)*. However, given that only five individuals in our study had a known history of anorexia, a follow up study would be needed to specifically evaluate the role of anorexia in our recurrent BSI models in a larger sample size. Problematic or prolonged low energy availability (LEA) from an eating disorder or disordered eating can lead to Relative Energy Deficiency in Sport (REDs), a syndrome in which bone quality and health are often impaired *(58)*. LEA is major risk factor for BSI occurrence *(59)*, so our finding of a history of anorexia being a positive predictor of subsequent BSI is not surprising. While no participants had a known current eating disorder throughout the study, it is plausible that they still had subclinical disordered eating or that there are lasting effects of a past anorexia diagnosis on metabolic function or lifestyle. Therefore, in our study, a history of anorexia may be supplemented in the models to simplify and improve the accuracy of the genetic programming models by replacing several individual proteins.

Our study achieved ∼97% accuracy in predicting additional BSIs using sPLS-DA and genetic programming models with primarily proteomic data and effectively identified candidate biomarkers in relatively small biological datasets. This highlights how high-resolution proteomic data can enhance the predictive power of models through the synergistic effects of standard machine learning models with the appropriate dataset. However, evaluating model performance solely on error rates may not fully assess effectiveness. Genetic programming suggests that increased complexity can lead to less generalizable models, making it crucial to examine model distributions within the ensemble. Ideally, ensembles should exhibit a ‘knee’ shape in their context plot, balancing error minimization and complexity. In our study, this ‘knee’ configuration was absent in models relying solely on clinical metrics, indicating inefficiencies and potential overfitting. Conversely, proteomic models showed lower error rates and retained the ‘knee’ shape. This demonstrates that integrating detailed biomolecular profiles enhances the robustness and accuracy of predictive models in clinical applications.

Our study has several limitations. While our results identified significant correlations between specific proteins and recurrent BSIs, establishing causal relationships would require further investigation. Multivariate models based on high dimensional, low sample size datasets are common in clinical research but face challenges like possible overfitting and increased complexity, complicating validation and interpretation. We sought to overcome these limitations by using both sPLS-DA and genetic programming models to explore both linear and non-linear relationships in the data as well as independent models with different assumptions. While most features presented as linear relationships in our datasets, there were subsets of proteomic data that were identified as non-linear relationships in genetic programming model ensembles, validating that both linear and non-linear relationships should be explored.

Our results were also limited to the demographics of the study group, which includes relatively healthy, young female athletes with diagnosed BSI as well as a skewed distribution of individuals. Although our study demonstrated clear clustering between the study groups, we lack a test population to fully validate the identified candidate biomarkers for subsequent BSIs. Additionally, we lacked an injury-free control group, which would have aided in understanding the protein levels regarding health or disease. Nevertheless, our study identified 10 significant candidate biomarkers that appeared in 80% of the sPLS-DA models, including FAAA, Osteopontin, and Trypsin-2, with a predictive accuracy of 95 ± 0.02% validated through leave-one-out cross-validation. Time-course differential expression analysis highlighted 112 differentially expressed proteins linked to pathways of increased fibrin clot formation and decreased immune signaling in individuals with additional BSIs. Our data highlight differences based on subsequent BSI risk in a relatively homogeneous study population (young, female athletes), adding to the strength of these findings as they relate to additional BSI risk in our study population.

The clinical implications of our study include the identification of (1) candidate biomarkers of subsequent BSIs in young, female athletes, (2) pathway signatures of immunosuppression and increased blood clotting with subsequent BSIs, and (3) an innovative approach to identifying and validating candidate biomarkers in relatively small clinical study populations. Further studies are required to prospectively validate FAAA, OPN, and Trypsin-2 as biomarkers of subsequent BSIs, as well as their relationship in a broader study population. Additionally, the relationship between functional immunosuppression and increased blood clotting should be further explored in athletes with BSI.

In conclusion, this study presents a robust statistical workflow, enhanced by machine learning, to analyze high-resolution proteomic data, paving the way for advancements in reinjury risk assessment. Our study underscores a critical gap in sports medicine: clinical metrics alone are insufficient to predict risk of recurrent BSIs, leading to potential discrepancies in determining appropriate rehabilitation and return to activity protocols. Our data reveal that specific protein signatures within three weeks of an initial BSI uniquely identify individuals at risk for subsequent BSI within 52 weeks. While biomarker discovery in small datasets is challenging, advances in proteomic technologies and machine learning can help isolate critical proteins that differentiate those with single BSIs from those at higher risk of additional BSI. Through multi-modeling, subsampling, and cross-validation, we consistently identified candidate biomarkers, reinforcing their potential as early indicators of additional BSIs. The network analysis of these proteins offers deeper insights into the biological mechanisms underlying subsequent BSI risk. These findings not only enhance our understanding of the pathophysiology of BSIs, but also suggest new avenues for preventive strategies and personalized medicine in sports, with the potential to reduce the incidence and impact of recurrent injuries.

## MATERIALS AND METHODS

### Study Design

The design of the clinical study was previously published *(11)*. In brief, local community recreational female runners who had been diagnosed with a BSI of MRI Fredericson grade 2-4/4 *(9)* were enrolled in the study and monitored over 52 weeks. Inclusion criteria included a minimum of 4 hours/week of self-reported weightbearing exercise during the 6 months prior to injury. Exclusion criteria included known medical conditions that would affect bone health (e.g., current eating disorder, hyperparathyroidism, celiac disease). Exclusion also included medications known to affect bone health (e.g., oral steroids, bisphosphonates, lithium). Enrollment attrition is summarized in Fig. 1, with loss due to disinterest (2 participants), diagnosis > 3 weeks (2 participants), lack of physical activity (1 participants), and dropped participation (2 participants). Four participants missed more than 1 visit (Fig. 1). Timepoints include enrollment (within 3 weeks of BSI diagnosis), 6, 12, 24, and 52 weeks post-enrollment. Participants were monitored for activity levels, bone density (DXA and HR-pQCT), clinical standard-of-care labs, and development of additional BSIs. Over the course of the study, 10 participants developed a 2^nd^ BSI, one of whom developed a 3^rd^ BSI (Table 1). The remaining 20 participants went on to heal and return to activity uneventfully. Following this study, we re-analyzed serum from all 30 final participants to retrospectively determine the proteomic signatures between those with a single vs. additional BSI. Proteomic data are all newly generated data and all analyses are new, including those with clinical data that have been previously published *(11)*.

To overcome the small clinical sample size, multiple machine learning algorithms were applied to determine robust proteomic signatures in those with an additional BSI compared to those with a single BSI. These models included sPLS-DA combined with non-parametric machine learning models: CatBoost *(60)*, logistic regression *(61)*, and random forest *(62)*. LOOCV was applied to sPLS-DA models to provide robust sub-sampling of the data during model building to ensure no single individual was skewing the protein signatures. Finally, protein signatures were analyzed for key biological pathway signatures to glean insights into the systemic dysregulation in those with additional BSIs. This study was approved by the institutional review boards of participating institutions and informed consent was obtained prior to participation.

### Dataset

The clinical study was previously published *(11)* and was restricted to the clinical and HR-pQCT data. New analyses of the serum proteins were completed and the subsequent bioinformatic approach combined the prior and new data. The current dataset consisted of 30 individuals, 10 of whom experienced a new BSI event during the 1 year of follow-up. For each participant, the serum was analyzed with a custom panel of 1,500 proteins by the SOMAscan (SomaLogic, Boulder, CO, USA) (Data File S2). Detailed methods for the SOMAscan assay have been previously published *(63)*. In brief, aptamer-based protein capture is utilized to multiplex the analysis and quantify protein levels in a microarray-based technology.

Initially, raw clinical data from the original study *(11)* were meticulously reviewed to discern which metrics were relevant. We excluded some variables, such as types and frequency of exercise and height, which were deemed extraneous and/or noisy for modeling BSI risk. Additionally, to maintain consistency across the data, categorical variables, such as contraceptive use, were standardized as categorical variables. A comprehensive workflow was then created to preprocess the data and create data structures amenable to machine learning approaches.

### Proteomic Panel Selection & SomaLogic Assay

Previous literature was initially examined to identify key proteins of interest. This led to the curation of a list containing 1,500 target proteins, each selected for its association with relevant physiologic categories from pre-fabricated panels provided by SomaLogic, such as ‘Cardiovascular Disease’, ‘Inflammation and Immune Response’, ‘Metabolic Disease’, and ‘Oncology’, as well as identified proteins of interest from literature *(64–66)* (full list in Data File S2). To efficiently align these proteins with the naming convention used by SomaLogic, a specialized Python program was developed. This program automatically translates gene names, as cited in prior research, into UniProt IDs. Consequently, a custom panel featuring these 1,500 relevant proteins was created, providing a focused and comprehensive proteomic assay for our study.

Following custom panel creation, serum samples from clotted blood were supplied to SomaLogic for aptamer-based analysis. Samples were obtained from 30 individuals from the prior BSI study *(11)*. SomaLogic assay has been described previously. In brief, samples are analyzed with custom slow off-rate aptamers designed for up to 7,000 human proteins. Concurrent analysis of the custom 1,500 protein panel is done from 55 uL of serum and samples are normalized with SomaLogic’s custom and robust internal and plate-to-plate normalization methods *(67)*. Standard mathematical normalization techniques (log2 and mean centering) were applied before further processing. A subset of samples were flagged (6/145 samples) because the samples failed to hybridize in the initial step and were re-run successfully. Flagged samples can occur with bubble formation or other technical difficulties during assay processing.

### Data Preprocessing

Optimal data preprocessing was identified by comparing the methods: standardization, variance stabilization normalization (VSN), quantile, linear-regression, and mean-centered approaches, each of which are widely accepted preprocessing methods. Resulting box-plot distributions were compared for equitable distribution of the data and standardization emerged as the most effective normalization technique. Transformation was similarly scrutinized, with log2, log10, and box-cox evaluated. The log2 transformation was selected because it stabilized the variance of high intensities but increased the variance at low intensities, allowing for more uniform evaluations *(68)*. Standardization and log2 transformations were applied to all datasets.

### Machine Learning Sparse Partial Least Square Discrimination Analysis (sPL-DA)

Clinical and proteomic data were compiled for analysis using custom Python and R scripts designed for feature reduction and multivariate modeling. Sparse Partial Least Square Discriminant Analysis (sPLS-DA) was conducted in R (R 4.4.1) using the MetaboAnalystR 4.0 *(69)* package. Prior to analysis, the data were preprocessed as described above.

The sPLS-DA method was utilized to reduce dimensionality while retaining the most informative features across clinical, proteomic, and combined datasets. Three components were computed, with each retaining 10 latent variables (LVs), which maximized group separation. An orthogonal rotation was applied in the component space to align the axis with the largest group distinction, thereby ensuring that the selected collection of LVs captured the most meaningful variation in the data. Score plots were evaluated and components with the largest degree of separation between ‘additional BSI’ and ‘single BSI’ were selected for further analysis.

To validate the robustness of the sPLS-DA model and minimize undue influence by single individuals with our relatively low sample size (n = 30), a feature stability test was performed. Here, one individual was randomly removed from the dataset and models were recorded. This was repeated 10 times. Models were then compared for the most frequently appearing features. Stable features were identified if present in over 80% of models during the 10 iterations. Only stable features were input into the final machine learning models.

Machine learning models were applied to the stable features including non-parametric machine learning models: CatBoost *(60)*, logistic regression *(61)*, and random forest *(62)*. These models were chosen because they do not assume a specific distribution of the data *(70)*, making them ideal for complex clinical and proteomic dataset. Feature selection was performed for each model with subsequent LOOCV, ensuring the most consistent and reliable features were evaluated. This cross-validation approach provided a rigorous evaluation of the model’s predictive capability, addressing the challenges posed by our dataset’s high dimensionality and limited sample size. Model performance was evaluated by ROC curves, accuracy, sensitivity, and specificity across the models.

### Nonlinear Multivariate Modeling with Genetic Programming

Multivariate, linear and nonlinear regression models were generated using Evolved Analytics DataModeler software (Version 9.7) in the SymbolicRegression package. Our modeled outcome was binary as “additional BSI” or “single BSI”. Preprocessed data were uploaded as “clinical”, “proteomic”, or “combined” (clinical + proteomic) datasets. Total models generated were 3,020 models for “clinical”, 2,435 models for “proteomic”, and 3,225 models for “combined”, each over 3 rounds of independent modeling at ten iterations each resulting in 30 total rounds of modeling. Models were then filtered to the “fittest” models, which represent ∼50% of models that satisfied < 80 complexity score and < 0.2 square error rate within the “knee” of the Pareto front. This reduced models to 65 for “clinical, 130 for “proteomic” and 258 for “combined” models. Model ensembles were generated and analyzed using the VariablePresence and CreateModelEnsemble functions, which represented the diversity of the filtered models for both complexity and square error rate. Model ensemble statistics are presented in the paper for the 10-model ensemble for “clinical”, 10-model ensemble for “proteomic”, and 11-model ensemble for “combined”.

### Differential Expression Analysis and Gene Set Enrichment Analysis

ExpressAnalyst package *(22)* based on the *limma* package *(23)* in R (R 4.4.1) was utilized for the differential expression analysis (DEA). Data was log2 transformed and mean-centered. Cutoff criteria included a fold change (FC) > 0.5. A Benjamini-Hochberg false discovery rate (FDR) adjusted p-value < 0.05 was applied. Full data can be found in Data File S5. Time was used as a blocking factor for the differential expression analysis. Gene Set Enrichment Analysis (GSEA) was used to map the statistical likelihood that sets of identified proteins relate to biological pathways based on the assumption that the measured proteins would map to gene databases of known interaction networks. This approach was used because protein interaction networks are less well developed for high-resolution proteomics and is common for this data type *(71)*. An FDR ≤ 0.05 and *p* < 0.05 was also used for the GSEA. A heatmap of the average protein values, z-scored across timepoints, was used to display the proteins corresponding to the top up- and down-regulated biological pathways.

### Statistical Analysis

Data are reported as mean ± SEM for time-course analysis of individual proteins. Statistical analysis was performed for time-dependent variation using a linear mixed effect model, *lme4* package *(72)*, in R (R 4.4.1) for time series analysis of select proteins with a Tukey’s post-hoc analysis for contrast results of “additional BSI” v. “single BSI”, specifically for key proteins in the sPLS-DA and genetic programming models. Statistical significance was determined at *p* < 0.05 and FDR ≤ 0.05 for all components of the study. sPLS-DA and Model ensemble information are presented in the respective sections above.

## Data Availability

All data produced in the present study are available upon reasonable request to the authors and will be published with the final peer-reviewed manuscript.

## List of Supplementary Materials

Figures S1-S5

Tables S1-S3

Data Files S1-S9

## Acknowledgments

We would like to acknowledge the clinical research team at Boston Children’s Hospital for efforts in patient recruitment, data collection, and management. We would also like to thank Dr. Mark Kotanchek for assistance with the DataModeler software, Dr. Dan Lowd for assistance with machine learning models, Dr. Clay Small and Dr. Karl Romanowicz for bioinformatic discussions, as well as Dr. Bill Cresko for reviewing the biostatistics, machine learning, and manuscript prior to submission. The opinions or assertions contained herein are the private views of the author(s) and are not to be construed as official or as reflecting the views of the Army, the Department of Defense, National Institute of Health, or the U.S. Government.

The opinions or assertions contained herein are the private views of the authors and are not to be construed as official or reflecting the views of the U.S. Army or the Department of Defense. Any citations of commercial organizations and trade names in this report do not constitute an official U.S. Army, Department of Defense endorsement of approval of the products or services of these organizations. This paper has been approved for public release with unlimited distribution.

Artificial intelligence (AI) assisted technologies (ChatGPT-4, GitHub Copilot leveraging ChatGPT4o) were used to aid in code revision and debugging as well as revision of text used in the manuscript. All code was evaluated and checked to ensure accuracy and is available on GitHub. Throughout the manuscript, text was reviewed after prompts such as “revise” and “condense” were used to aid in clarification of the text and reducing word count. The authors take full responsibility for both the coding and written content of this manuscript.

## Funding

United States Department of Defense, Defense Health Program, and Joint Program Committee (W811XWH-15-C-0024) (PI: Bouxsein ML);

Wu Tsai Human Performance Alliance (Multi-PIs: Ackerman KE, Guldberg RE);

NIH-NIDCR, K99DE033689 (PI: Romanowicz GE);

Knight Campus Undergraduate Scholarship (Dinh)

## Author contributions

Conceptualization: GER, KP, ED, IH, KL, KEA, MLB, REG

Methodology: GER, KP, ED, IH, KL, REG

Investigation: GER, KP, ED, IH, JMH, KEA

Visualization: GER, ED, IH

Funding acquisition: GER, MLB, KEA, REG

Project administration: GER, KP, MLB, REG

Supervision: GER, KP, MLB, REG

Writing – original draft: GER, ED

Writing – review & editing: GER, KP, ED, IH, KL, JMH, KEA, MLB, REG

## Data and materials availability

All data are available in the main text or the supplementary materials. Code relating to the analysis is available on GitHub: https://github.com/GuldbergLab/BSI-Study-2024

