## Supplemental Materials for "Deciphering Risk of Recurrent Bone Stress Injury in Female Runners Using Serum Proteomics Analysis and Predictive Models"

### List of Supplementary Materials

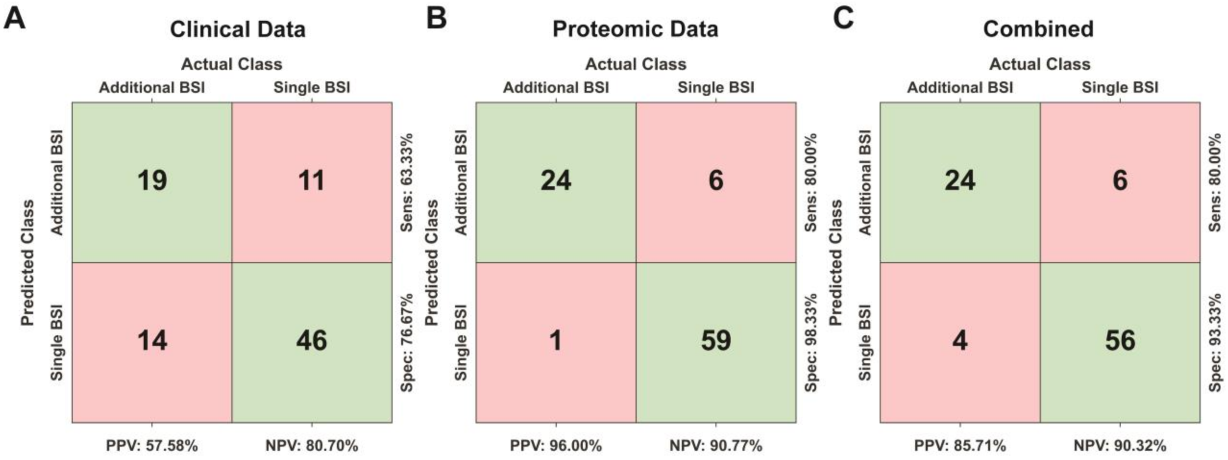


**Fig. S1.** Confusion matrix from the resulting sparse partial least square discrimination analysis (sPLS-DA) models for (A) "clinical data", (B) "proteomic data", and (C) "combined".

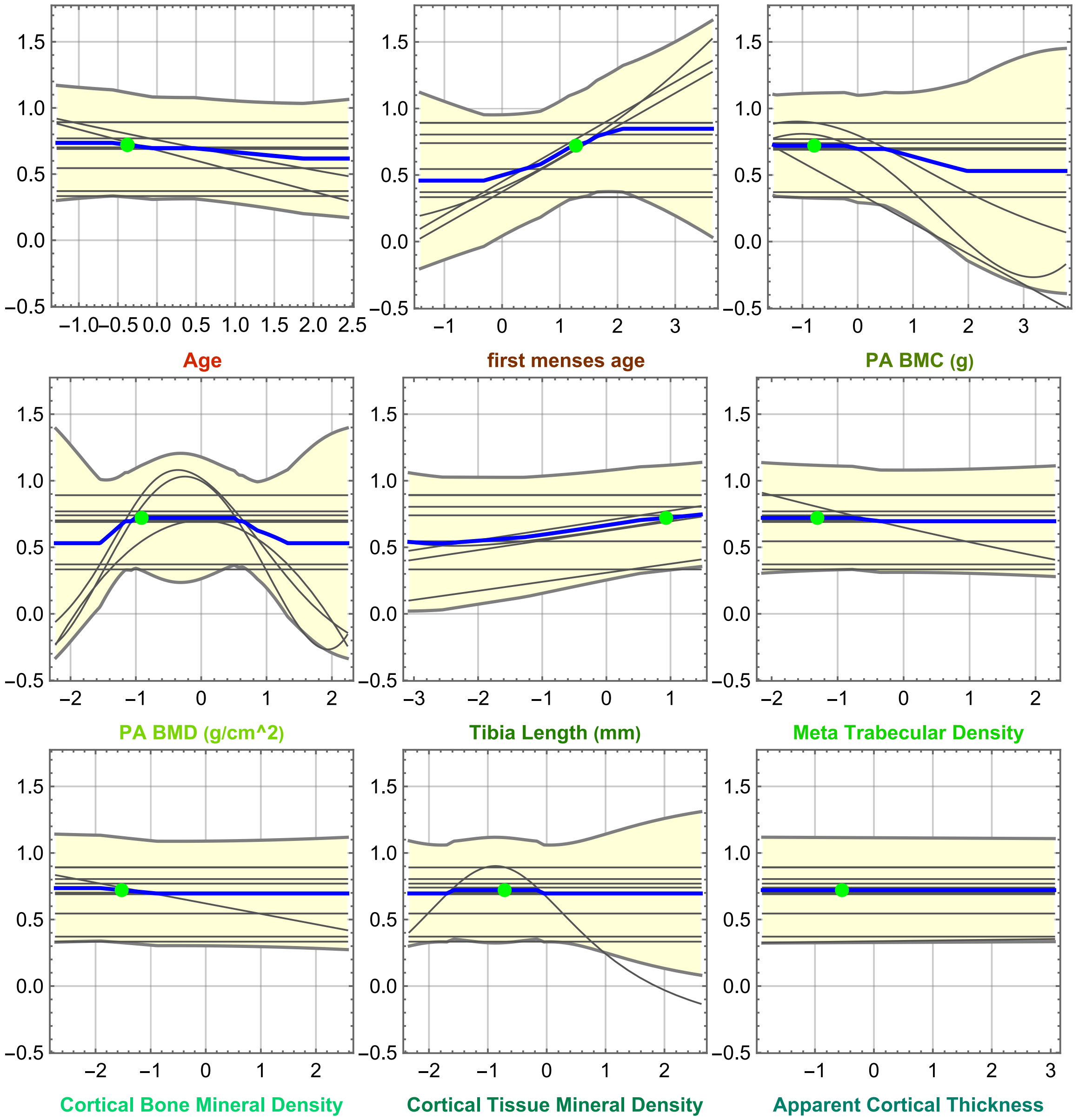


**Fig. S2.** Response plots from genetic programming model ensembles based on clinical data. The blue line represents the mean value of the predictive ensemble, grey lines represent individual models and the yellow band represents the ensemble variance for each variable. Most variables present a linear relationship with subsets of variables and models having non-linear relationships including PA-BMC and PA-BMD.

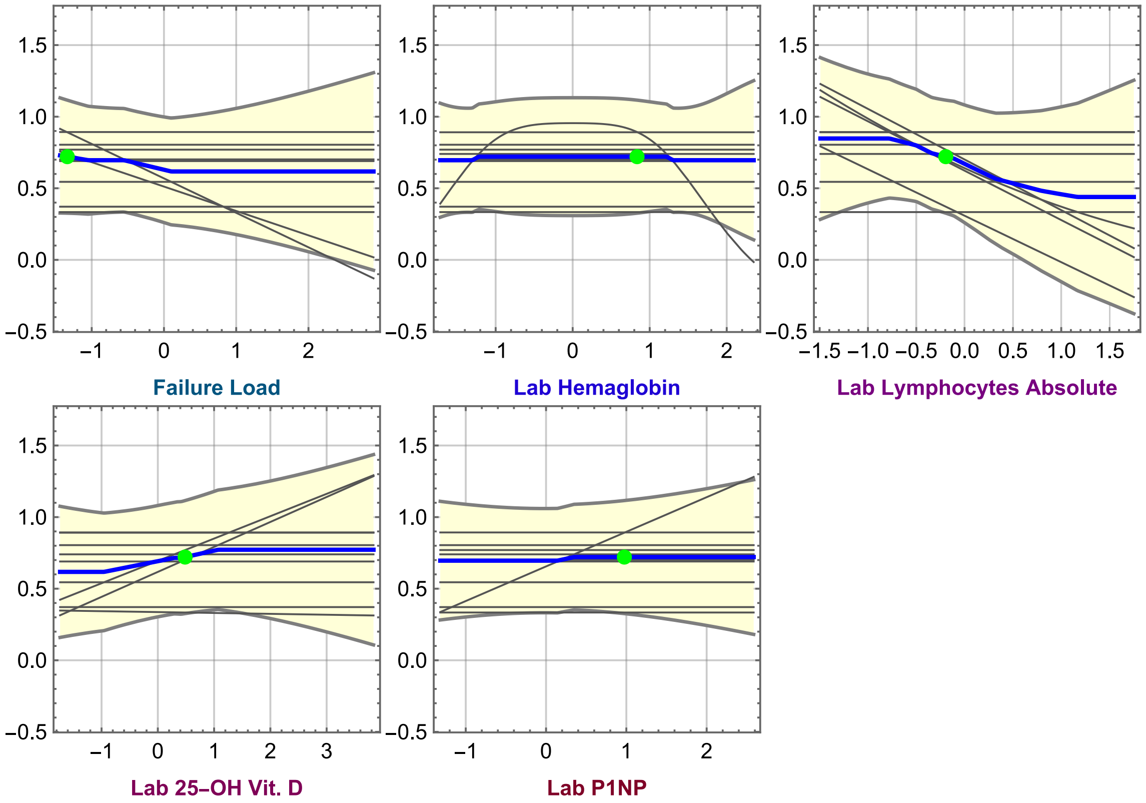


**Fig. S2.** (Continued)

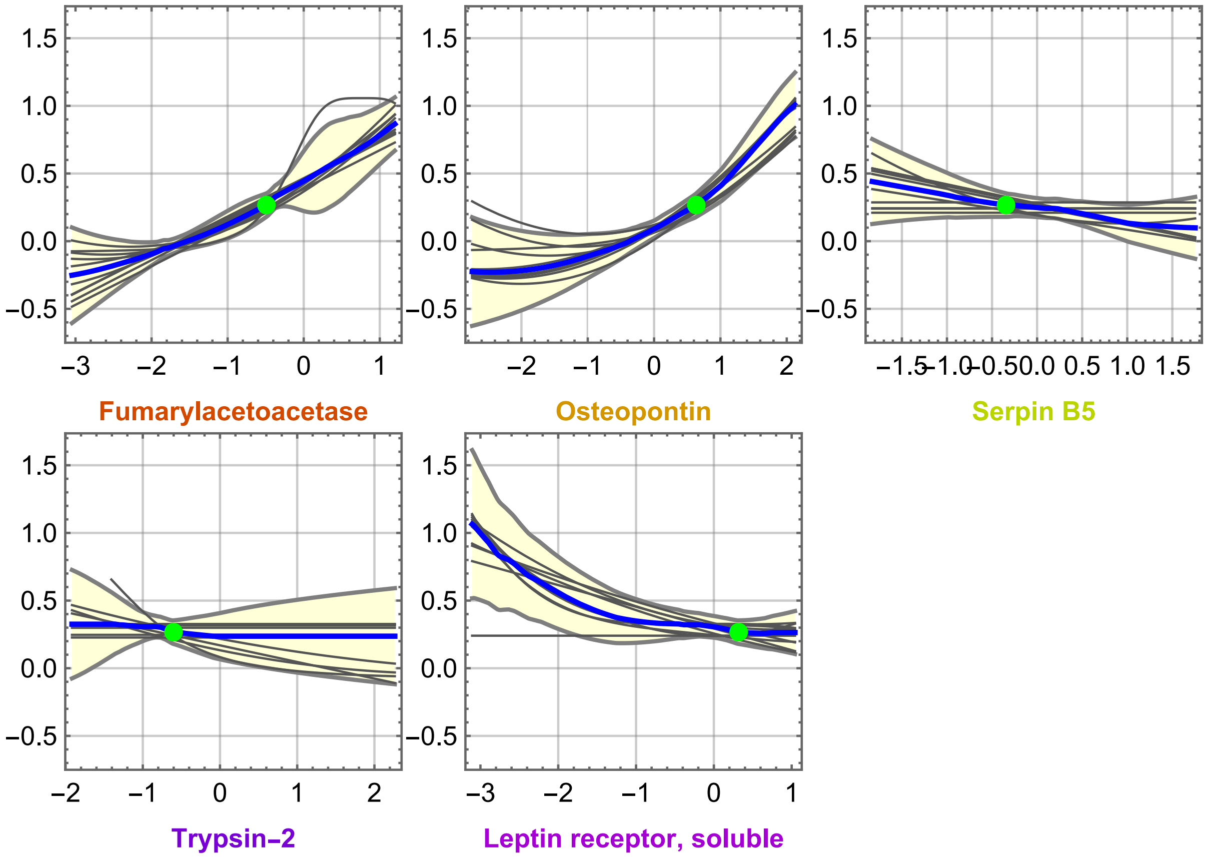


**Fig. S3.** Response plots from genetic programming model ensembles based on clinical data. The blue line represents the mean value of the predictive ensemble, grey lines represent individual models and the yellow band represents the ensemble variance for each variable. Resultant models were primarily linear with non-linear models for OPN and LEPR.

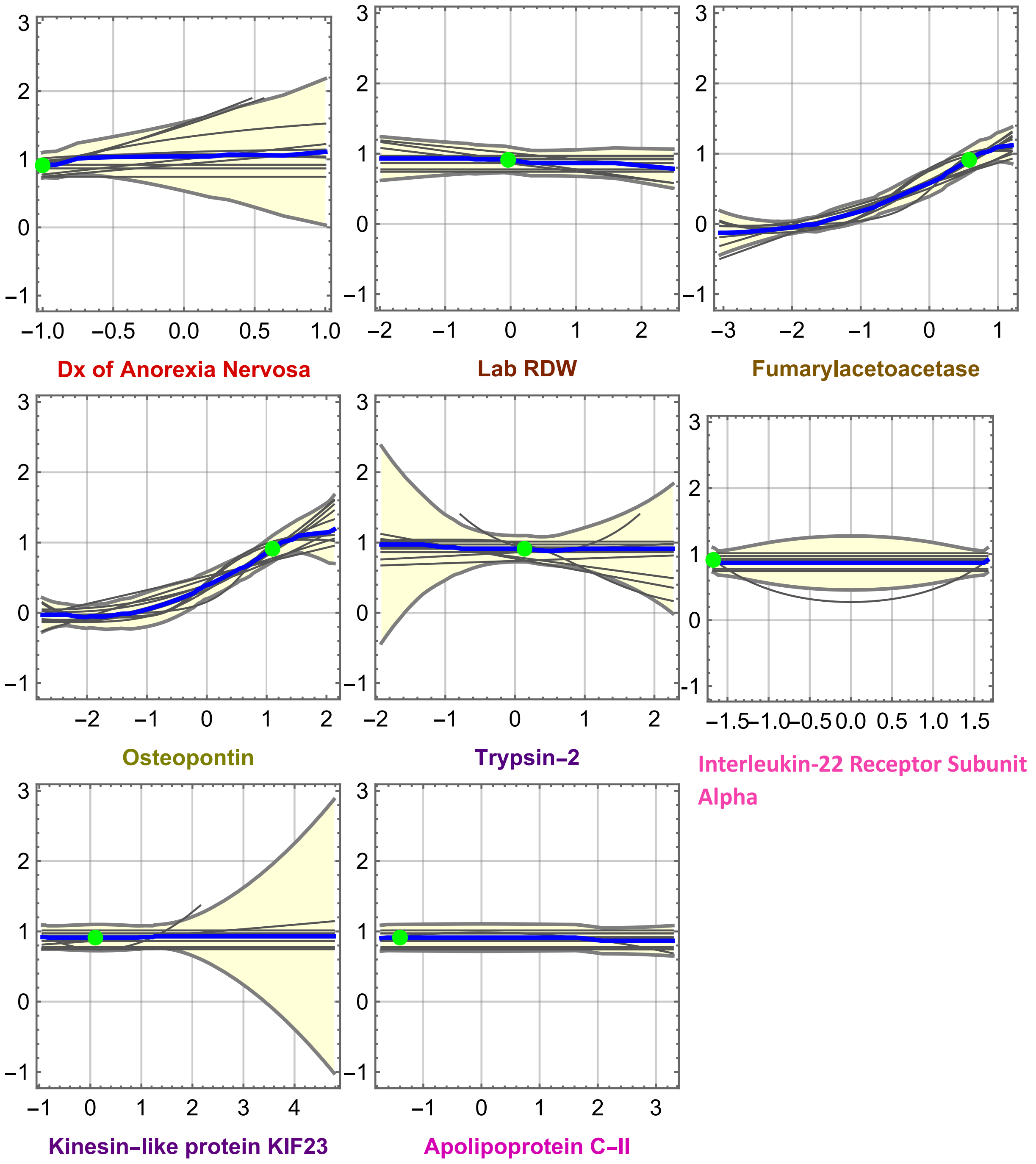


**Fig. S3.** Response plots from genetic programming model ensembles based on clinical data. The blue line represents the mean value of the predictive ensemble, grey lines represent individual models and the yellow band represents the ensemble variance for each variable. Resultant models were primarily linear with non-linear models for FAAA and OPN.

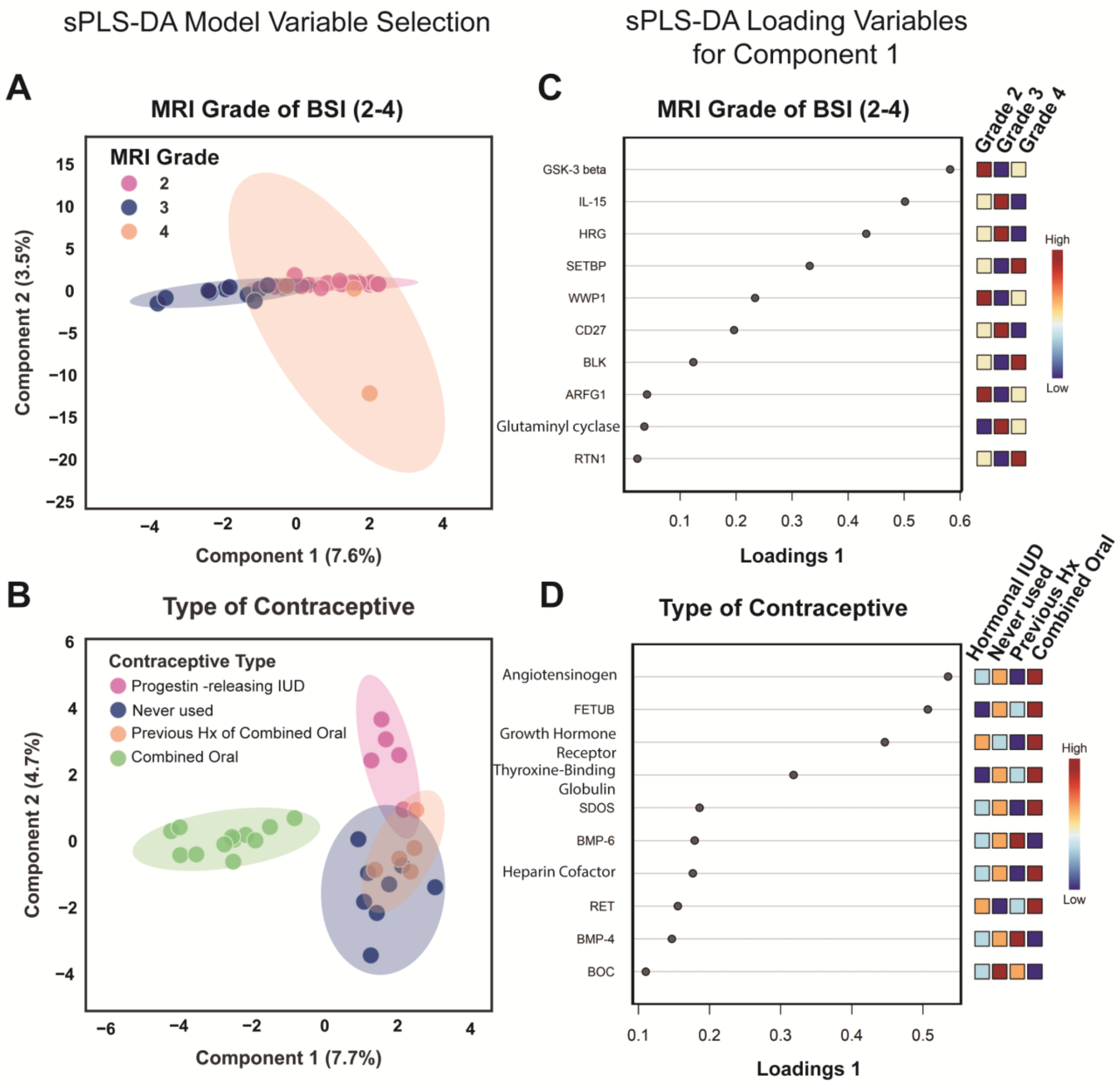


**Fig. S4.** Analysis of proteomic signatures relating type of contraceptive and bone stress injury (BSI) magnetic resonance imaging (MRI) grade to determine if key proteins from BSI recurrence models overlap with proteins present in contraceptive type or MRI grade features. (A) Sparse partial least squares discrimination analysis (sPLS-DA) analysis summary for MRI grade of the BSI showing negligible separation between groups with only some spread along component one. (B) sPLS-DA for type of contraceptive showing separation primarily along component 1 between those using combined oral contraceptive pills compared to progestin-releasing hormonal intrauterine device (IUD) or non-users of contraceptive ('previous history of combined oral' or 'never used' for contraceptive). Shaded regions represent the 95% confidence interval. (C&D) Summary of component one loading variables which are all distinct from models corresponding to BSI recurrence, indicating that it is unlikely that the protein signatures present in BSI models are due to type of contraceptive or MRI grade. Full list of protein names in Data File S7.

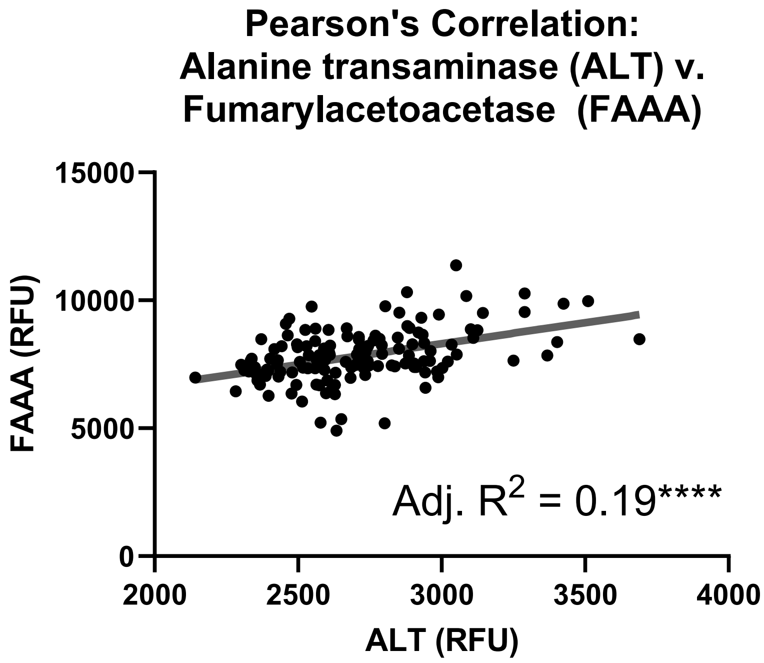


**Fig. S5.** Correlation of alanine transaminase (ALT) and fumarylacetoacetase (FAAA) from the proteomic data reveals a significant, positive correlation (Pearson's correlation, Adj. $R^2 = 0.19$, ****$p < 0.0001$). All units are relative fluorescence units (RFUs).

**Table S1.** Model ensemble for the 10 selected models from the genetic programming algorithm based on clinical data alone

| | Complexity | $1-R^2$ | Variables | Function |
|---|---|---|---|---|
| 1 | 14 | 0.620 | Tibia Length (mm)<br>Lab Lymphocytes Absolute | $0.31 - 0.35 \cdot$ Lab Lymphocytes Absolute $\cdot$ Tibia Length (mm) |
| 2 | 14 | 0.999 | Apparent Cortical Thickness<br>Lab 25-OH Vit. D | $0.34 + (1.13 \cdot 10^{-2}) \cdot$ Apparent Cortical Thickness $\cdot$ Lab 25-OH Vit. D |
| 3 | 19 | 0.588 | Failure Load<br>Lab P1NP | $0.33 - 0.24 \cdot$ Failure Load $+ 0.24 \cdot$ Lab P1NP |
| 4 | 22 | 0.346 | first menses age<br>Tibia Length (mm)<br>Lab lymphocytes Absolute | $0.31 + 0.25 \cdot$ first menses age<br>$-0.27 \cdot$ Lab Lymphocytes Absolute $\cdot$ Tibia Length (mm) |
| 5 | 23 | 0.657 | PA BMC (g)<br>PA BMC $(g/cm^2)$ | $0.52 - 0.23 \cdot$ PA BMC (g) $- 0.19 \cdot$ PA BMC $(g/cm^2)$ |
| 6 | 27 | 0.668 | Age<br>Cortical Bone Mineral Density<br>Failure Load | $0.33 - 0.16 \cdot$ Age $- (7.82 \cdot 10^{-1}) \cdot$ Cortical Bone Mineral Density<br>$-0.16 \cdot$ Failure Load |
| 7 | 30 | 0.237 | first menses age<br>Tibia Length (mm)<br>Lab Lymphocytes Absolute<br>Lab 25-OH Vit. D | $0.31 + 0.25 \cdot$ first menses age $+ 0.16 \cdot$ Lab 25-OH Vit. D<br>$-0.38 \cdot$ Lab Lymphocytes Absolute $\cdot$ Tibia Length (mm) |
| 8 | 99 | 0.307 | Age<br>PA BMC (g)<br>PA BMC $(g/cm^2)$<br>Meta Trabecular Density | $-0.27 \cdot (3.54 \cdot 10^{-3}) \cdot (-16.25 +$ Age $+$ Meta Trabecular Density<br>$+2 \cdot$ PA BMC (g) $+$ PA BMC $(g)^2 + 2 \cdot$ PA BMC $(g/cm^2)$<br>$+4 \cdot$ PA BMC $(g/cm^2)^2)^2$ |

Continued on next page

**Table S1.** Model ensemble for the 10 selected models from the genetic programming algorithm based on clinical data alone (Continued)

| | | | | |
|---|---|---|---|---|
| 9 | 121 | 0.205 | first menses age<br>Tibia Length (mm)<br>Lab Lymphocytes Absolute<br>Lab 25-OH Vit. D | $0.22 + 0.21 \cdot \text{first menses age} + 0.17 \cdot \text{Lab 25-OH Vit. D} - (3.4 \cdot 10^{-2}) \cdot \text{Lab Lymphocytes Absolute} - 0.39 \cdot \text{Lab Lymphocytes Absolute} \cdot \text{Tibia Length (mm)} + (1.99 \cdot 10^{-2}) \cdot \text{Tibia Length (mm)}^2 + (3.17 \cdot 10^{-2}) \cdot (\text{first menses age} + \frac{\text{Lab Lymphocytes Absolute} \cdot \text{Tibia Length (mm)}}{4} + \text{Lab Lymphocytes Absolute} \cdot \text{Tibia Length (mm)}^2)^2$ |
| 10 | 205 | 0.140 | PA BMC (g)<br>PA BMC $(g/cm^2)$<br>Cortical Tissue Mineral Density<br>Lab Hemaglobin | See footnote[1] |

[1] $$97.08 + \frac{487.61}{-5 + \frac{1}{-12.20 + \text{Lab Hemaglobin}^2 3 \cdot \text{PA BMC } (g/cm^2) - 5 \cdot \text{PA BMC } (g/cm^2)^2 - (1 + 2 \cdot \text{Cortical Tissue Mineral Density} + \text{Lab Hemaglobin}^2 + \text{PA BMC (g)} + \text{PA BMC } (g/cm^2)^2)^2}}$$

**Table S2.** Model ensemble for the 10 selected models from the genetic programming algorithm based on proteomic data alone.

| | Complexity | $1-R^2$ | Variables | Function |
|---|---|---|---|---|
| 1 | 51 | 0.115 | Fumarylacetoacetase<br>Leptin receptor, soluble<br>Osteopontin<br>Serpin B5 | $0.19 + 0.28 \cdot \text{Fumarylacetoacetase} - (2.67 \cdot 10^{-2}) \cdot \text{Leptin receptor, soluble}^3 + 0.31 \cdot \text{Osteopontin} + (6.56 \cdot 10^{-2}) \cdot \text{Osteopontin}^2 - 0.14 \cdot \text{Serpin B5}$ |
| 2 | 59 | 0.111 | Fumarylacetoacetase<br>Leptin receptor, soluble<br>Osteopontin<br>Serpin B5 | $0.19 + 0.29 \cdot \text{Fumarylacetoacetase} - (8.52 \cdot 10^{-2}) \cdot \text{Leptin receptor, soluble} + (5.16 \cdot 10^{-2}) \cdot \text{Leptin receptor, soluble}^2 + 0.31 \cdot \text{Osteopontin} + (6.75 \cdot 10^{-2}) \cdot \text{Osteopontin}^2 - 0.13 \cdot \text{Serpin B5}$ |
| 3 | 62 | 0.118 | Fumarylacetoacetase<br>Leptin receptor, soluble<br>Osteopontin<br>Trypsin-2 | $-10.36 - 1.49 \cdot \text{Fumarylacetoacetase} - 0.16 \cdot \text{Leptin receptor, soluble} - 1.57 \cdot \text{Osteopontin} + (7.57 \cdot 10^{-2})(11.83 + \text{Fumarylacetoacetase} + \text{Osteopontin})^2 - 0.12 \cdot \text{Trypsin-2}$ |
| 4 | 63 | 0.102 | Fumarylacetoacetase<br>Leptin receptor, soluble<br>Osteopontin<br>Serpin B5 | $8.85 \cdot 10^{-2} + 0.29 \cdot \text{Fumarylacetoacetase} + (8.36 \cdot 10^{-2}) \cdot \text{Leptin receptor, soluble}^2 + 0.32 \cdot \text{Osteopontin} + (7.98 \cdot 10^{-2}) \cdot \text{Osteopontin}^2 - 0.13 \cdot \text{Serpin B5} + (6.50 \cdot 10^{-2}) \cdot \text{Serpin B5}^2$ |
| 5 | 67 | 0.114 | Fumarylacetoacetase<br>Leptin receptor, soluble<br>Osteopontin<br>Serpin B5 | $0.20 + 0.30 \cdot \text{Fumarylacetoacetase} + (1.50 \cdot 10^{-3}) \cdot \text{Fumarylacetoacetase}^3 - (2.66 \cdot 10^{-3}) \cdot \text{Leptin receptor, soluble}^3 + 0.30 \cdot \text{Osteopontin} + (6.38 \cdot 10^{-3}) \cdot \text{Osteopontin}^2 - 0.14 \cdot \text{Serpin B5}$ |
| 6 | 67 | 0.118 | Fumarylacetoacetase<br>Leptin receptor, soluble<br>Osteopontin<br>Serpin B5 | $0.19 + 0.28 \cdot \text{Fumarylacetoacetase} + (2.74 \cdot 10^{-4}) \cdot \text{Leptin receptor, soluble}^2 + (8.72 \cdot 10^{-3}) \cdot \text{Leptin receptor, soluble}^4 + 0.30 \cdot \text{Osteopontin} + (6.50 \cdot 10^{-2}) \cdot \text{Osteopontin}^2 - 0.14 \cdot \text{Serpin B5}$ |
| 7 | 73 | 0.101 | Fumarylacetoacetase<br>Leptin receptor, soluble<br>Osteopontin<br>Serpin B5 | $0.17 + 0.29 \cdot \text{Fumarylacetoacetase} - (5.57 \cdot 10^{-2}) \cdot \text{Leptin receptor, soluble} + (7.30 \cdot 10^{-3}) \cdot \text{Leptin receptor, soluble}^4 + 0.28 \cdot \text{Osteopontin} + (6.62 \cdot 10^{-2})(\text{Fumarylacetoacetase} + \text{Osteopontin})^2 - 0.11 \cdot \text{Serpin B5}$ |

Continued on next page

**Table S2.** Model ensemble for the 10 selected models from the genetic programming algorithm based on proteomic data alone. (Continued)

| | | | | |
|---|---|---|---|---|
| 8 | 76 | 0.112 | Fumarylacetoacetase<br>Leptin receptor, soluble<br>Osteopontin<br>Trypsin-2 | $-(4.09 \cdot 10^{-2}) + (1.96 \cdot 10^{-2})(3 + 2 \cdot \text{Fumarylacetoacetase}$<br>$-\text{Leptin receptor, soluble} + 2 \cdot \text{Osteopontin} - \text{Trypsin-2})^2$ |
| 9 | 92 | 0.107 | Fumarylacetoacetase<br>Leptin receptor, soluble<br>Osteopontin<br>Trypsin-2 | $-0.19 + (1.16 \cdot 10^{-2}) \cdot \text{Leptin receptor, soluble} + (1.17 \cdot 10^{-2})(5.92$<br>$+2 \cdot \text{Fumarylacetoacetase} - \text{Leptin receptor, soluble}$<br>$+2 \cdot \text{Osteopontin} - \text{Trypsin-2})^2 + (3.06 \cdot 10^{-2}) \cdot \text{Trypsin-2}$ |
| 10 | 93 | 0.068 | Fumarylacetoacetase<br>Osteopontin<br>Trypsin-2 | $-(8.01 \cdot 10^{-2}) + \frac{7.96}{7+0.44(-2.78+2 \cdot \text{Fumarylacetoacetase}+\text{Osteopontin}-\text{Trypsin-2})^4}$<br>See footnote[2] for full formatting |

[2] $-(8.01 \cdot 10^{-2}) + \dfrac{7.96}{7 + 0.44(-2.78 + 2 \cdot \text{Fumarylacetoacetase} + \text{Osteopontin} - \text{Trypsin-2})^4}$

**Table S3.** Model ensemble for the 11 selected models from the genetic programming algorithm based on combined clinical and proteomic data.

| | **Complexity** | $1 - R^2$ | **Variables** | **Function** |
|---|---|---|---|---|
| 1 | 35 | 0.153 | History of Anorexia<br>Fumarylacetoacetase<br>Osteopontin<br>Trypsin-2 | $0.52 + 0.22 \cdot \text{History of Anorexia} + 0.35 \cdot \text{Fumarylacetoacetase}$<br>$+0.37 \cdot \text{Osteopontin} - 0.13 \cdot \text{Trypsin-2}$ |
| 2 | 43 | 0.105 | Lab RDW<br>History of Anorexia<br>Fumarylacetoacetase<br>Osteopontin | $-0.13 - 0.11 \cdot \text{Lab RDW}$<br>$+(8.31 \cdot 10^{-2})(2.78 + \text{History of Anorexia}$<br>$+\text{Fumarylacetoacetase} + \text{Osteopontin})^2$ |
| 3 | 51 | 0.083 | History of Anorexia<br>Lab RDW<br>Fumarylacetoacetase<br>Osteopontin | $-0.21 - (9.59 \cdot 10^{-2}) \cdot \text{History of Anorexia} - 0.13 \cdot \text{Lab RDW}$<br>$+(8.55 \cdot 10^{-2})(2.78 + \text{History of Anorexia}$<br>$+\text{Fumarylacetoacetase} + \text{Osteopontin})^2$ |
| 4 | 53 | 0.068 | History of Anorexia<br>Fumarylacetoacetase<br>Osteopontin | $1.17 - \frac{11.96}{9.69+(2+\text{History of Anorexia}+\text{Fumarylacetoacetase}+\text{Osteopontin})^4}$<br>See footnote[3] for full formatting |
| 5 | 58 | 0.130 | History of Anorexia<br>Fumarylacetoacetase<br>Osteopontin<br>Trypsin-2 | $0.43 + 0.18 \cdot \text{History of Anorexia} + 0.30 \cdot \text{Fumarylacetoacetase}$<br>$+0.27 \cdot \text{Osteopontin} + (5.08 \cdot 10^{-2}) \cdot \text{Osteopontin}^2 - 0.15$<br>$\cdot\text{Trypsin-2} - (5.20 \cdot 10^{-2}) \cdot \text{Fumarylacetoacetase} \cdot \text{Trypsin-2}$ |
| 6 | 63 | 0.114 | Kinesin-like protein KIF23<br>Osteopontin<br>Fumarylacetoacetase | $-(8.15 \cdot 10^{-2}) - (1.97 \cdot 10^{-2}) \cdot \text{Kinesin-like protein KIF23}$<br>$-(3.95 \cdot 10^{-2}) \cdot \text{Osteopontin} + (7.29 \cdot 10^{-2})(2$<br>$+\text{Fumarylacetoacetase} + \text{Osteopontin}$<br>$+\text{Kinesin-like protein KIF23})^2$ |

Continued on next page

**Table S3.** Model ensemble for the 11 selected models from the genetic programming algorithm based on combined clinical and proteomic data. (Continued)

| | | | | |
|---|---|---|---|---|
| 7 | 65 | 0.097 | Apolipoprotein C-II<br>History of Anorexia<br>Fumarylacetoacetase<br>Osteopontin | $1.80-(2.74\cdot10^{-2})\cdot\text{Apolipoprotein C-II}^2 - \frac{264.38}{140.47+(3+\text{History of Anorexia}+\text{Fumarylacetoacetase}+\text{Osteopontin})^4}$<br>See footnote[4] for full formatting |
| 8 | 69 | 0.140 | Fumarylacetoacetase<br>Kinesin-like protein KIF23<br>Osteopontin<br>Trypsin-2 | $.29+0.38\cdot\text{Fumarylacetoacetase}+(4.86\cdot10^{-2})\cdot\text{Fumarylacetoacetase}^2-0.10\cdot\text{Kinesin-like protein KIF23}+0.20\cdot\text{Osteopontin}+0.13\cdot\text{Kinesin-like protein KIF23}^2\cdot\text{Osteopontin}^2+0.34\cdot\text{Kinesin-like protein KIF23}\cdot\text{Trypsin-2}$ |
| 9 | 69 | 0.029 | History of Anorexia<br>Lab RDW<br>Fumarylacetoacetase<br>Osteopontin | $1.14-(7.42\cdot10^{-2})\cdot\text{History of Anorexia}-(9.00\cdot10^{-2})\cdot\text{Lab RDW}-\frac{12.27}{9.69+(2+\text{History of Anorexia}+\text{Fumarylacetoacetase}+\text{Osteopontin})^2}$<br>See footnote[4] for full formatting |
| 10 | 94 | 0.004 | History of Anorexia<br>Fumarylacetoacetase<br>Osteopontin<br>Trypsin-2 | $-(3.36\cdot10^{-2})+\frac{30.08}{28+(-3.41+\text{History of Anorexia}+2\cdot\text{Fumarylacetoacetase}+2\cdot\text{Osteopontin}-\text{Trypsin-2})^4}$<br>See footnote[5] for full formatting |
| 11 | 95 | 0.052 | Fumarylacetoacetase<br>Osteopontin<br>Trypsin-2<br>Interleukin-22 receptor subunit alpha-2 | $-(8.79\cdot10^{-2})-(1.84\cdot10^{-2})\cdot\text{Fumarylacetoacetase}+(2.02\cdot10^{-2})(1.06+2\cdot\text{Fumarylacetoacetase}+\text{Interleukin-22 receptor subunit alpha-2}^2+2\cdot\text{Osteopontin}-\text{Trypsin-2}+\text{Trypsin-2}^2)^2$ |

[3] $1.17-\frac{11.96}{9.69+(2+\text{History of Anorexia}+\text{Fumarylacetoacetase}+\text{Osteopontin})^4}$

[4] $1.14-(7.42\cdot10^{-2})\cdot\text{History of Anorexia}-(9.00\cdot10^{-2})\cdot\text{Lab RDW}-\frac{12.27}{9.69+(2+\text{History of Anorexia}+\text{Fumarylacetoacetase}+\text{Osteopontin})^2}$

[5] $-(3.36\cdot10^{-2})+\frac{30.08}{28+(-3.41+\text{History of Anorexia}+2\cdot\text{Fumarylacetoacetase}+2\cdot\text{Osteopontin}-\text{Trypsin-2})^4}$

**Data File S1**. Summary of the variables included in the clinical data collected from our prior study (Popp, et al., 2019). Clinical data was used in the sparse partial least square discrimination analysis (sPLS-DA) as well as the genetic programming models. Missing values are noted per group.

**Data File S2.** Summary of the custom protein panel analyzed using the Somalogic aptamer based proteomic array. The entirety of the proteomic data was used in the sparse partial least square discrimination analysis (sPLS-DA) as well as the genetic programming models. Subsets of data were utilized for the differential expression analysis and gene set enrichment analysis and are designated where appropriate.

**Data File S3.** Resulting latent variables for the sparse partial least square discrimination analysis (sPLS-DA) including a tab for "clinical data", "proteomic data", and "combined data".

**Data File S4.** Feature stability analysis for the sparse partial least square discrimination analysis (sPLS-DA) models to determine the frequency of features selected across models when one individual was randomly removed over 10 iterations. Data is presented for "clinical data", "proteomic data", and "combined data". Variables in bold met the >80% frequency to be categorized as a "stable feature" and included in the machine learning models.

**Data File S5.** Summary of the 112 significantly different proteins identified in the differential protein expression analysis for individuals when blocking for time ($\log_2$ FC > 0.5, FDR < 0.05, $p < 0.05$).

**Data File S6.** Summary of the gene set enrichment analysis utilizing the proteins identified in the differential expression analysis.

**Data File S7.** Resulting latent variables for the sparse partial least square discrimination analysis (sPLS-DA) including a tab for "MRI Grade of BSI (2-4)" and "Type of Contraceptive".

**Data File S8**. Raw data for participants from the SomaScan assay. All values are in relative fluorescence units (RFUs).

**Data File S9**. Raw data for participants from the clinical dataset. Values are included in the file and subcategorized based on the acquisition method.
